# Positive deviance for promoting dual-method contraceptive use among women in Uganda: a cluster randomized controlled trial

**DOI:** 10.1101/2020.10.03.20206169

**Authors:** Hodaka Kosugi, Akira Shibanuma, Junko Kiriya, Ken Ing Cherng Ong, Stephen Mucunguzi, Conrad Muzoora, Masamine Jimba

**Affiliations:** Department of Community and Global Health, Graduate School of Medicine, The University of Tokyo, Tokyo, Japan; UNICEF Uganda Country Office, Kampala, Uganda; Department of Internal Medicine, Mbarara University of Science and Technology, Mbarara, Uganda

**Author notes:** Corresponding Author: Professor Masamine Jimba; Department of Community and Global Health, Graduate School of Medicine, The University of Tokyo, 7-3-1 Hongo, Bunkyo-ku, Tokyo 113-0033, Japan.

## Abstract

**Background:** Dual-method contraceptive use, or using condoms with highly effective contraceptives, is effective at preventing both unintended pregnancies and HIV infections. Although it remains uncommon among women in long-term relationships in sub-Saharan Africa, some do practice it. The positive deviance approach aims to promulgate practices of such individuals to other members in the community. We examined the effects of a positive deviance intervention on the dual-method use among married or in-union women.

**Methods:** We conducted a cluster randomized controlled trial in 20 health facilities in Mbarara District, Uganda, with 960 women aged 18–49 years. The intervention was a combination of clinic- and phone-based counseling and a participatory workshop, which was developed based on the qualitative study of women practicing the dual-method use in the study area. The control group received regular clinic-based counseling and health-related messages via phone. We assessed dual-method contraceptive use at the last sexual intercourse and its consistent use at two, four, six, and eight months after enrollment.

**Findings:** More women in the intervention group reported dual-method contraceptive use at the last sexual intercourse at two months (AOR = 4.29; 95% CI: 2.12–8.69; p < 0.001) and at eight months (AOR = 2.19; 95% CI: 1.07–4.48; p = 0.032) than in the control group. Consistent dual-method contraceptive use was also more prevalent in the intervention group at two months (AOR = 13.71; 95% CI: 3.59–52.43; p < 0.001), and the intervention effect remained at four, six, and eight months.

**Conclusion:** Dual-method contraceptive use increased significantly among women in the intervention group. The positive deviance intervention can be a potential option for promoting the dual-method use among women in long-term relationships in Uganda.

**Key questions:** 

**What is already known about this topic?:**

- Dual-method contraceptive use is incredible for preventing both unintended pregnancies and HIV infections but remains uncommon among women in long-term relationships in sub-Saharan Africa.
- The positive deviance approach aims to promulgate behaviors of individuals who have achieved rare success to other members in the community and has the potential to achieve sustainable behavioral change.

**What are the new findings?:**

- The proportion of women practicing dual-method contraception at the last sexual intercourse increased from 8.5% at baseline to 42.6% at two months after enrollment in the intervention group.
- Over 15% and 11% of women in the intervention group reported consistent dual-method contraceptive use at two and eight months, respectively.
- Between the intervention and control groups, the significant difference was detected for their consistent dual-method contraceptive use during the eight-month follow-up period.

**What do the new findings imply?:**

- The positive deviance intervention is effective in reducing the dual risk of unintended pregnancies and HIV infections through promoting dual-method contraceptive use among women in Mbarara District, Uganda.
- The positive deviance approach can help women to uptake and adhere to dual-method contraceptive use by disseminating the local solutions found among women already practicing it in the community.

## Introduction

Unintended pregnancy and human immunodeficiency virus (HIV) infection are major public health concerns in sub-Saharan Africa (SSA). It is estimated that 29% of pregnancies are unintended, whereas women account for 59% of the 980,000 new HIV infections among adults in SSA every year.^1,2^ Sexual intercourse is a major route of HIV transmission, and a significant gender disparity in HIV infection begins when women reach reproductive age.^3^ In SSA, therefore, women of reproductive age bear the dual burden of unintended pregnancies and HIV.

Dual-method contraceptive use has been proposed as an effective strategy for preventing unintended pregnancies and sexually transmitted infections (STIs), including HIV.^4^ Dual-method contraception is defined as the use of a highly effective contraceptive (HEC) (e.g., injectables, implants, and oral contraceptive pills, intrauterine devices, and sterilization) in combination with a barrier method, such as male or female condoms.^4^ Despite the high incidence rate of HIV, it is not commonly practiced in SSA, especially among women in long-term relationships.^4,5^ For instance, only 3.8% of married women in Zimbabwe used dual-method contraception with their partners.^5^ Furthermore, women in stable relationships tend to prioritize HECs over condoms and are less likely to use condoms with HECs.^6–8^

Although the majority of women understand that using condoms is critical for preventing HIV/STIs, they do not practice it.^9^ Using condoms in long-term relationships is often considered unacceptable or awkward, especially for women using HECs.^8^ Marital sexual intercourse becomes one of the major routes of HIV infection because of inconsistent or no condom use in SSA.^10^

Several studies examined interventions for promoting dual-method contraceptive use.^4^ However, few showed a significant effect on the dual-method use, and their impact was often unsustainable.^11^ To our knowledge, the only intervention that demonstrated a continued effect on the dual-method use over six months was a combination of case management and peer leadership programs among adolescents in the United States of America (USA).^12^ In SSA, conditional lottery incentives increased dual-method use among South African women at three months but not at six months after the intervention.^13^ Effectiveness of behavioral change interventions on the dual-method use among married or in-union women remains lacking.

The positive deviance approach is based on the premise that there are people who solve problems while many of their peers do not.^14^ This approach seeks unique behaviors of such exceptional people (positive deviants or PDs) and disseminates these behaviors to the whole community through community-led and peer-based interventions.^14,15^ We previously conducted a qualitative study to examine the unique behaviors of PDs (i.e., women using dual-method with marital or in-union partners) in Mbarara District, Uganda.^16^ These PDs successfully practiced dual-method contraception by initiating discussions, educating their partners on sexual risks and condom use, and obtaining condoms.^16^ In this study, we examined the effectiveness of an intervention developed based on those findings to promote dual-method contraceptive use among women in the same area.

## Methods

### Study design and settings

A cluster randomized controlled trial was conducted for eight months (November 2019 to July 2020) in Mbarara District in Southwestern Uganda. The protocol of the trial has been previously published.^17^ The prevalence of HIV is geographically diverse in Uganda, and the Southwestern region has one of the highest prevalence rates of HIV at 7.9% among adults. This rate is higher among women (9.3%) than men (6.3%).^18^ An estimated 32% and 2% of married or in-union women use HECs and condoms, respectively.^19^ All public health facilities provide HECs and male condoms free of charge. Male condoms are also available for purchase at pharmacies and markets.^16^

### Study participants and enrollment

Twenty public health facilities were selected out of 48 in Mbarara District. To recruit a sufficient number of participants, all health facilities at the sub-county level or above were selected followed by health facilities at the parish level, which had a high number of outpatients. These facilities included one general hospital, three county-level health centers, 11 sub-county-level health centers, and five parish-level health centers. Among them, seven facilities were located in urban areas.^20^

The inclusion criteria were women (i) aged 18 to 49 years, (ii) having had sexual intercourse in the last three months, (iii) using HECs, and who (iv) desire to avoid pregnancy for 12 months from recruitment, (v) have a husband or live-in sexual partner, and (vi) have access to a valid phone number. The exclusion criteria were women who were (i) pregnant, (ii) infertile for other reasons, and (iii) had been using condoms consistently with an HEC in the last two months before the recruitment. The sample size of 960 was calculated based on the effect size of 2.43 reported in a dual-method intervention trial in the USA, considering an intraclass correlation coefficient of 0.006 and a 26% dropout rate.^11,12,21^ The power of the study was set at 80%, and the significance level was set at 5%.

Female research assistants recruited women who visited the family planning sections of the selected health facilities. They approached every third woman after selecting the first woman purposively to inform the opportunity to participate in the study. If a woman was interested, they confirmed HEC use with her family planning client record card and asked questions to verify eligibility. The process was repeated until the required sample size was reached. All women received an information sheet for their partners, which included the objectives of the study and contact information of the researchers for notifications in the case of conflict with their partners.

### Randomization and masking

The 20 health facilities were stratified based on their level and urban or rural status. They were then randomized to the intervention or control group with a 1:1 allocation ratio. An independent researcher who was uninvolved in the data collection and analysis carried out the allocation using computer-generated random sequences. Blinding was not feasible in this study. However, the research assistants who were engaged in the outcome assessment were blinded to the allocation.

### Intervention

The intervention was developed based on the results of the preliminary study of nine PDs conducted in Mbarara District, Uganda in October 2019. The PDs were identified by screening 150 women using HECs at five health facilities. In-depth interviews were conducted with the PDs. Thematic analysis was then performed using the positive deviance framework to identify the unique behaviors associated with dual-method contraceptive use. The findings of the study have been published.^16^

Out of the nine PDs, four joined the intervention as peer counselors, whereas the other five were unable to participate due to other commitments. The four PDs demonstrated dual-method contraceptive use at least three months before the screening. The mean age of the four PDs was 29.8 years (standard deviation [SD] 6.0 years). The researchers (HK and SM) initially developed the intervention based on the preliminary findings. The PDs were then invited to four meetings. In the first meeting, two female research assistants explained the positive deviance approach and facilitated a discussion among the PDs to share their experience on how they started the dual-method use. In the following meetings with the PDs, they facilitated discussions on how to promote the dual-method use and necessary improvements to the intervention. Several recommendations from the PDs were incorporated into the intervention, such as providing a handout to enable women to share topics learned with their partners and effective communication skills, which were practiced through role-play.

Table 1 summarizes the intervention, which combined clinic- and phone-based counseling and a participatory workshop, to disseminate the unique practices of the PDs.^16^ After the baseline interview on the day of enrollment, women received counseling focusing on dual-method contraception in addition to regular family planning counseling. Trained research assistants delivered the counseling for approximately 20 to 30 minutes. Women received the handout used during the counseling developed either in English or Runyankore and were encouraged to initiate discussions on the dual-method use with their partners. The handout included several quotes from the PDs.

**Table 1.** Overview of the intervention.

| Training setting | Duration | Topics covered |
| --- | --- | --- |
| Clinic-based counseling | 20–30 minutes | 1. Comparing family planning methods<br>2. HIV/STI risk<br>3. Means to avoid HIV/STIs<br>4. Introduction and demonstration of male condoms<br>5. Effective communication with partners<br>6. Information about the workshop |
| One-day workshop at a health facility | 5 hours | 1. Introduction of family planning methods<br>2. Means to avoid unintended pregnancies<br>3. Introduction of HIV/STI risk<br>4. Means to avoid HIV/STIs<br>5. Group discussion 1: Let's consider your HIV/STI risk<br>6. Practice of condom use<br>7. Experience of four PDs<br>8. Role-play exercises: Effective communication with partners<br>– Initiating discussions on condom use<br>– How to persuade partners<br>– How to avoid conflicts<br>9. Group Dissuasion 2: Recapitulate takeaway messages<br>– Why is dual-method contraceptive use important?<br>– What are the barriers to dual-method contraceptive use, and how can you overcome them? |
| Bimonthly phone-based counseling | 15–30 minutes each | 1. Brief health message:<br>– Family planning methods (at 3 months)<br>– Means to avoid HIV/STIs (at 5 months)<br>– General facts about HIV/STIs (at 7 months)<br>2. Counseling tailored to individual participants' situation and needs |

HIV: human immunodeficiency virus; STI: sexually transmitted infection; PD: positive deviant

After two weeks of enrollment, women were invited for a one-day participatory learning workshop at the same health facility where they were recruited. Participation in the workshop was voluntary. The four PDs facilitated the workshop with support from one research assistant. It included role-play exercises to enable women to acquire successful communication skills of the PDs, practice of male condom use, and group discussions on the dual risk of unintended pregnancies and HIV/STI infection from their partners.

In addition, women in the intervention group received a bi-monthly telephone counseling call from the PDs three times (i.e., three, five, and seven months after enrollment). It aimed to confirm women’s dual-method status and challenges, provide reminders regarding the risk of unintended pregnancies and HIV/STIs, and strengthen their capacity to communicate with their partners. In addition, the call included brief health education messages on family planning and HIV/STI based on an existing tool.^22^ Each PD provided the same women with counseling during each call to build rapport and ensure effective counseling. Each counseling lasted for 15 to 30 minutes. The PDs maintained a counseling record and held a group meeting after each counseling period to reflect on the problems of women and advice given. The research assistants facilitated such meetings and answered questions from the PDs.

Women in the control group received family planning counseling, including condom use, from female research assistants for 10 to 20 minutes using the existing tool on the day of enrollment.^22^ However, this group of women did not receive the handout. Furthermore, the research assistants provided bimonthly health education three times (i.e., three, five, and seven months after enrollment) by phone. The topics were the same as those for the intervention group. Each call lasted for approximately ten minutes.

At the selected health facilities, condoms were provided for free regardless of group allocation. Before providing the intervention, the research assistants received a two-day training on the contents of the existing counseling tool. In addition, the four PDs received a one-day training on counseling and ethics, including the confidentiality of their clients. The PDs joined the intervention as volunteers but received 30,000 UGX (equivalent to 9 USD) per day when they engaged in the workshop and counseling to compensate for their time and transportation.

### Outcomes

The primary outcome was dual-method contraceptive use, which was defined as the application of a male or female condom along with an HEC, such as injectables, implants, intrauterine devices, pills, and female sterilization.^4^ It was measured in two timeframes: dual-method contraceptive use at the last sexual intercourse and its consistent use in the last two months before each follow-up. The former is easier for women to answer accurately than the latter, which requires to estimate the frequency of condom use in the past.^23^ Nevertheless, consistent dual-method contraceptive use is critical, given that condoms are often used inconsistently.^23^ Two questions regarding HEC use and the frequency of condom use were combined to measure consistent dual-method contraceptive use. The following question was posed for HEC use: “Apart from condoms, have you been using any other forms of protection against pregnancy during the past two months?” The frequency of condom use was asked with an item: “How often did you and your partner use a male or female condom during the past two months?” Women answered this question using a four-point scale “every time,” “almost every time,” “sometimes,” and “never.” Women using an HEC and a condom every time were considered practicing consistent dual-method contraceptive use.

The secondary outcome was communication about HIV/STI risk with partners in the last two months prior to each follow-up. This outcome was assessed using the following item: “Have you ever discussed HIV/STI risk with your husband/live-in sexual partner in the past two months?” Another secondary outcome was the self-reported incidence of pregnancy in the two months before each follow-up regardless of whether the pregnancy was intended or not. This outcome was assessed using the following questions: “Have you been told by a healthcare provider that you got pregnant for the first time in the past two months?”

In addition, the following information was collected at baseline: age, education, religion, employment, wealth index based on the availability of 18 household assets, number of children, respondent’s and partner’s pregnancy intention, history of unintended pregnancy, multiple sex partnership, type of HECs in use, respondent’s and partner’s HIV status, risk perception of HIV/STIs, HIV-related knowledge (HIV-KQ-18),^24^ condom use self-efficacy,^25^ and sexual relationship control power (the Sexual Relationship Power Scale).^26^ Several changes were made to the outcomes after the trial commenced. An outcome for STI incidence was omitted because we found that the reliability of self-reported STI incidence could be low among the participants during the data collection. Instead, the more measurable outcome of HIV/STI risk communication was added as a possible predictor of dual-method contraceptive use.

### Data collection

All research assistants received a two-day training on data collection and ethics before the baseline data collection. After enrollment, the research assistants interviewed women to identify their baseline characteristics using a pre-tested structured questionnaire. Each interview lasted approximately 30 to 45 minutes.

For outcome assessment, three female research assistants carried out follow-up phone calls bimonthly for eight months to assess the influence of the intervention on the primary and secondary outcomes (i.e., two, four, six, and eight months after enrollment). The participants received a text message reminding them to answer the next call or call back if they missed the first call. The assistants called each participant up to five times during each follow-up until they answered. The participants received incentives worth 20,000 UGX (equivalent to 6 USD) for their time after the baseline interview.

### Data analysis

Chi-squared tests and independent sample t-tests were performed to compare the general characteristics between the intervention and control groups at baseline and follow-up. Mixed-effects logistic regression analysis was performed to assess the effects of the intervention on the following outcomes: dual-method contraceptive use at the last sexual intercourse, consistent dual-method contraceptive use, communication about HIV/STI risk with partners, and the incidence of pregnancy in the two months before each follow-up. Unadjusted odds ratios (ORs) were first estimated by comparing between the control and intervention groups (Model 1). Then, in the main model (Model 2), the intervention effects were presented with adjusted odds ratios (AORs) for the interaction term (group × time) after controlling for cluster effects for all health facilities and the individuals. The AORs can be interpreted as the difference between the intervention and control groups in the outcome measures between baseline and each follow-up point. In the full model (Model 3), sociodemographic characteristics at baseline were controlled for in addition to the variables included in Model 2. For sensitivity analyses, attrition rates and reasons for dropout were compared between the intervention and control groups using Pearson’s chi-squared test. Moreover, differences in baseline characteristics were compared between women lost to follow-up and those who were reached. Analyses were conducted on an intention-to-treat basis. Significance level was set at 5%. STATA version 14 was used for data analyses.

### Ethics

Participation in this study was voluntary, and the participants provided written informed consent. The protocol was registered at UMIN-CTR Clinical Trial under identifier number UMIN000037065. The Consolidated Standards of Reporting Trials (CONSORT) checklist is available as Supplementary Table S1.

## Results

### Participant characteristics

Out of 1,956 women screened, 960 women were considered eligible for the trial and allocated to the intervention and control groups (Figure 1). Eighty women were recruited from each general hospital and county-level health center, whereas 40 women were recruited from each sub-county-level and parish-level health center. Of 480 women in the intervention group, 345 (71.9%) attended the one-day workshop. Moreover, 385 (80.2%), 361 (75.2%), and 369 (76.9%) received counseling at three, five, and eight months after enrollment, respectively. The response rate to the follow-up surveys ranged from 76.5% at two months to 82.3% at eight months. Women in the intervention group were more likely to respond at two months (79.8% vs 73.1%; p = 0.015) and four months (84.6% vs 79.4%; p = 0.036). The baseline characteristics were compared between women followed up and those lost to follow-up from each group (Supplementary Table S2).

**Figure 1.**
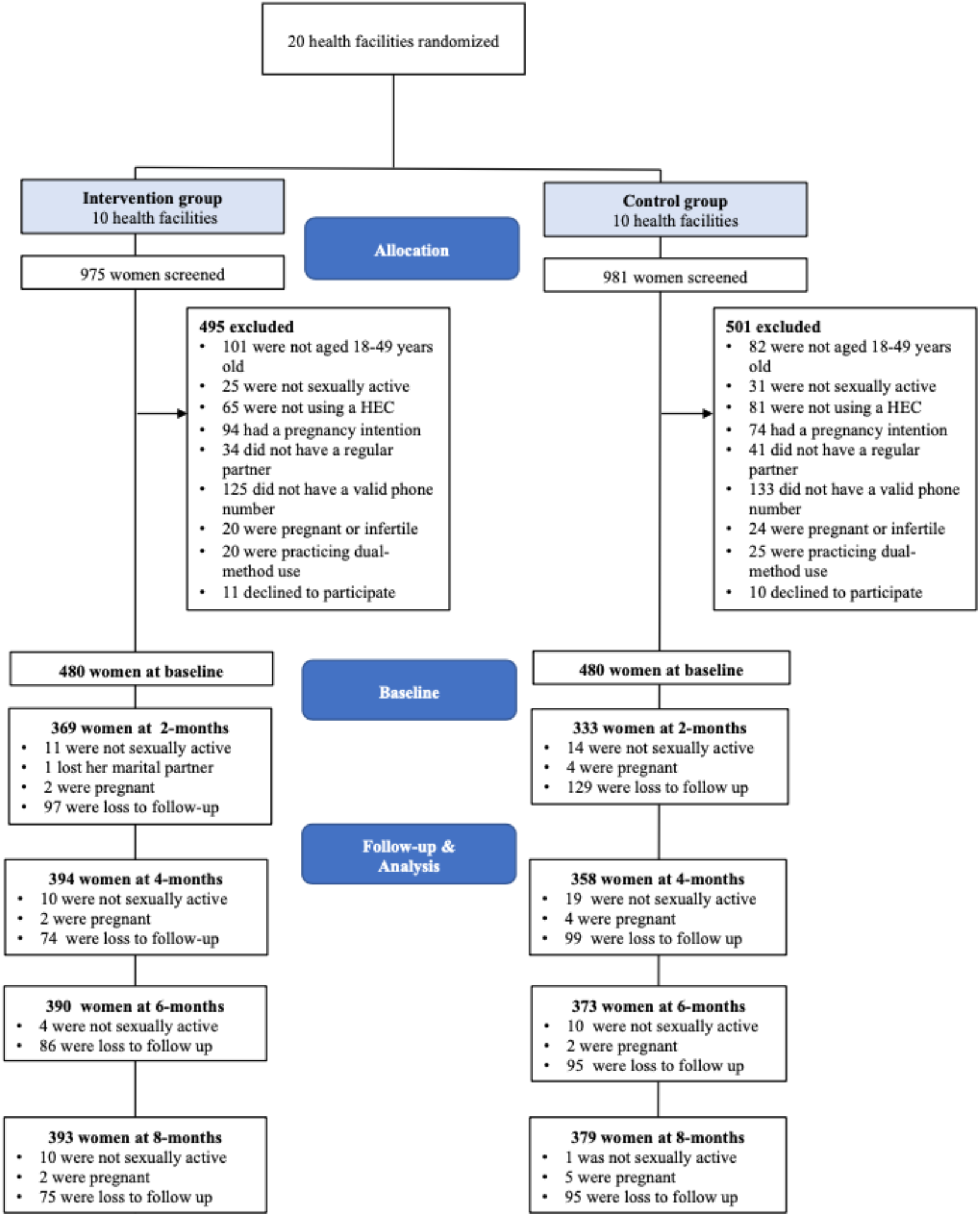
Flow of participants through the study.

Table 2 presents the sociodemographic characteristics of 960 women at baseline. The mean age was 30.1 (SD: 6.7) years. The mean number of children was three (SD: 1.8). Of 960 women, more than 70% completed primary education. In addition, 9% were HIV-positive, 7.6% had an HIV-positive partner, and 84.5% perceived a certain level of risk for HIV/STIs. Injectables were the most common HEC (51.9%), followed by implants (31.6%). Characteristics were similar for the intervention and control groups with a few slight imbalances. Specifically, women in the control group were more likely to have primary or higher education (75.6% vs. 69.8%; p = 0.042), be categorized into the rich quintile (37.7% vs. 28.3%; p = 0.008), and have less children (mean: 2.9 vs. 3.2; p = 0.041) and HIV-related knowledge (mean: 11.3 vs. 11.9; p < 0.001).

**Table 2.**
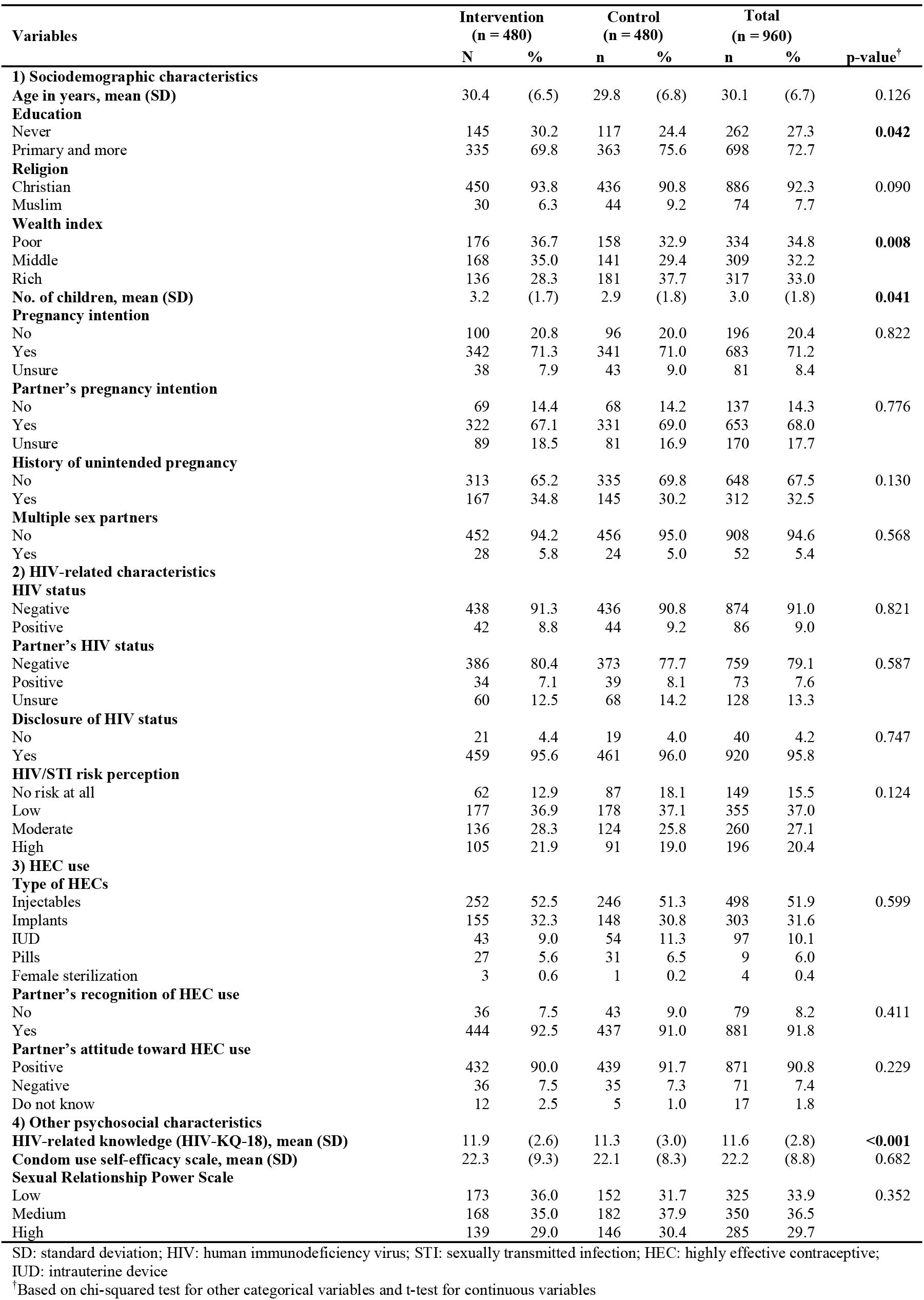
Characteristics of women at baseline by group (n = 960)

### Effect of the intervention

Table 3 presents the data of outcomes by group and time. More women in the intervention than in the control group used dual-method contraception at the last sexual intercourse and consistently at each follow-up point. These differences were largest at two months (dual-method contraceptive use at last sexual intercourse: 42.6% vs. 13.8%; p < 0.001; consistent dual-method contraceptive use: 15.5% vs. 1.5%; p < 0.001). The proportion of women practicing the dual-method decreased for both groups over time. More women discussed HIV/STI risk with their partners in the intervention than in the control group at each follow-up. The difference was also largest at the first follow-up (83.5% vs. 64.9%; p < 0.001). However, the incidence of pregnancy was not significantly different between the groups. Throughout the data collection period, 6 and 15 women became pregnant in the intervention and control groups, respectively. Notably, the result of the chi-squared test of the accumulated cases of pregnancies in eight months illustrated a significantly lower incidence of pregnancy in the intervention group (p = 0.047).

**Table 3.**
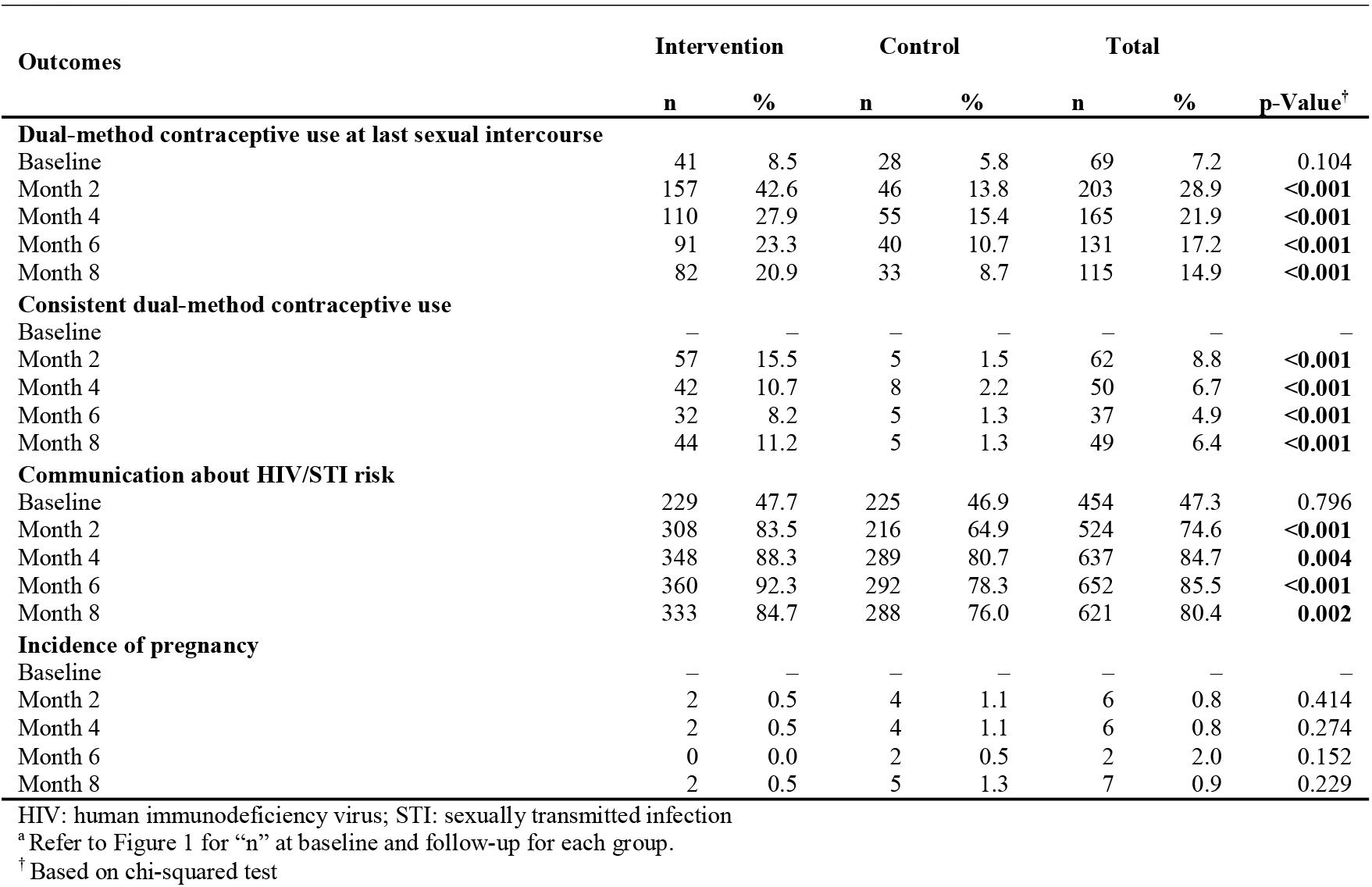
Dual-method contraceptive use, communication about HIV/STIs, and incidence of pregnancy by group and time

Table 4 illustrates the effects of the intervention on the primary and secondary outcomes at two, four, six, and eight months after enrollment. Model 2 indicates that more women in the intervention reported dual-method contraceptive use at the last sexual intercourse at two months (AOR = 4.29; 95% CI: 2.12–8.69; p < 0.001) than in the control group. The intervention group also reported more dual-method contraceptive use at the last sexual intercourse at four, six, and eight months, although the difference was statistically significant only at eight months. More women in the intervention group practiced consistent dual-method contraceptive use at two months (AOR = 13.71; 95% CI: 3.59–52.43; p < 0.001). The effect of the intervention remained statistically significant at four, six, and eight months. Moreover, more women in the intervention group reported communication with their partners regarding HIV/STI risk at two months (AOR = 2.70; 95% CI: 1.72–4.23, p < 0.001). The effect of intervention lasted throughout the follow-up period. However, the incidence of pregnancy was not significantly different between the groups throughout the follow-up period. The full model (Model 3) showed effect estimates similar to those reported in the main model. The complete results are provided in Supplementary Tables S3-S17.

**Table 4.** Effects of intervention on primary and secondary outcomes among women at two, four, six, and eight months after enrollment.

| Variables | Month 2 |  |  | Month 4 |  |  | Month 6 |  |  | Month 8 |  |  |
| --- | --- | --- | --- | --- | --- | --- | --- | --- | --- | --- | --- | --- |
|  | Model 1 | Model 2 | Model 3 | Model 1 | Model 2 | Model 3 | Model 1 | Model 2 | Model 3 | Model 1 | Model 2 | Model 3 |
|  | OR<br>(95% CI) | AOR<sup>a</sup><br>(95% CI) | AOR<sup>b</sup><br>(95% CI) | OR<br>(95% CI) | AOR<sup>a</sup><br>(95% CI) | AOR<sup>b</sup><br>(95% CI) | OR<br>(95% CI) | AOR<sup>a</sup><br>(95% CI) | AOR<sup>b</sup><br>(95% CI) | OR<br>(95% CI) | AOR<sup>a</sup><br>(95% CI) | AOR<sup>b</sup><br>(95% CI) |
| **Dual-method contraceptive use at last sexual intercourse** | 4.62***<br>(3.18–6.71) | 4.29***<br>(2.12–8.69) | 4.12***<br>(2.02–8.39) | 2.13***<br>(1.49–3.06) | 1.66<br>(0.84–3.30) | 1.66<br>(0.84–3.30) | 2.53***<br>(1.69–3.79) | 2.04<br>(1.00–4.17) | 2.03<br>(0.99–4.14) | 2.76***<br>(1.79–4.26) | 2.19*<br>(1.07–4.48) | 2.16*<br>(1.06–4.41) |
| **Consistent dual-method contraceptive use** | 11.98***<br>(4.74–30.29) | 13.71***<br>(3.59–52.43) | 14.53***<br>(3.63–58.13) | 5.22***<br>(2.42–11.28) | 6.28**<br>(2.01–19.60) | 6.30**<br>(2.20–18.03) | 6.58***<br>(2.53–17.07) | 7.80*<br>(1.22–49.73) | 8.04*<br>(1.17–55.08) | 9.43***<br>(3.70–24.06) | 9.97**<br>(2.11–47.15) | 10.7**<br>(2.03–56.64) |
| **Communication about HIV/STI risk** | 2.73***<br>(1.92–3.90) | 2.70***<br>(1.72–4.23) | 2.70***<br>(1.72–4.24) | 1.81**<br>(1.21–2.71) | 1.76*<br>(1.08–2.86) | 1.76*<br>(1.07–2.89) | 3.33*<br>(2.13–5.20) | 3.23***<br>(1.93–5.41) | 3.35***<br>(1.99–5.66) | 1.75**<br>(1.22–2.52) | 1.75*<br>(1.12–2.74) | 1.80*<br>(1.14–2.84) |
| **Incidence of pregnancy** | 0.69<br>(0.15–3.09) | 0.69<br>(0.15–4.69) | 1.27<br>(0.19–8.65) | 0.47<br>(0.09–2.57) | 0.47<br>(0.09–2.57) | 0.24<br>(0.00–16.42) | Perfect success |  |  | 0.38<br>(0.07–1.96) | 0.38<br>(0.07–2.19) | 0.40<br>(0.02–8.19) |

HIV: human immunodeficiency virus; STI: sexually transmitted infection
Note: The effect estimates are reported using odds ratio (OR) and adjusted odds ratio (AOR) from multiple logistic regression using the control group as the reference category.
\*\*\* p < 0.001.
\*\* p < 0.01.
\* p < 0.05.
<sup>a</sup> Adjusted for the cluster effect and individuals.
<sup>b</sup> Adjusted for cluster effect, individuals, age, education, religion, wealth index, number of children, pregnancy intention, partner's pregnancy intention, history of unintended pregnancy, multiple sex partnership, HEC methods, HIV status, partner's HIV status, HIV/STI risk perception, HIV-related knowledge, condom use self-efficacy, and sexual relationship control power.

## Discussion

The positive deviance intervention was effective in promoting dual-method contraceptive use and communication about HIV/STI risk among women using HECs in long-term relationships. However, no significant difference was observed in the incidence of pregnancy between the intervention and control groups. To the best of our knowledge, this is the first randomized controlled trial to demonstrate the effectiveness of an intervention to promote the dual-method use among married or in-union women in Uganda. The positive deviance approach can be a potential option for promoting dual-method contraceptive use by disseminating the unique behaviors of PDs associated with the dual-method use.

The positive deviance intervention increased the uptake and continued use of dual-method contraception among women. The study observed the largest difference in the dual-method use between the intervention and control groups at the two-month assessment, which was the closest time point to the baseline counseling and workshop. In the intervention group, 43% and 16% of women reported the dual-method use at the last sexual intercourse and its consistent use, respectively. The number of women using dual-method contraception decreased in the intervention and control groups over time, as observed in previous studies.^11^ Only 21% and 11% of women in the intervention group used the dual-method at the last sexual intercourse and consistently at the eight-month assessment, respectively. However, the significant difference between the groups remained during the follow-up period. The observed effect was consistent with a previous intervention study that combined case management and peer education program for adolescent girls in the USA. The intervention illustrated continued effects on the dual-method use at 12 and 24 months after enrollment.^12^ The peer leadership program aimed to foster prosocial interaction skills and supportive peer relationships among teenagers. The peer supporters were not PDs and provided with intensive standard training. Effective communication with partners on sexual health was one of the key topics covered in the sessions.^12^ Similar to this, the current intervention provided bimonthly counseling tailored to the participants’ individual needs. However, it was provided by the PDs who had overcome barriers to the dual-method use. Counseling by PDs may be an alternative strategy because it ensures adequate attention to the diverse issues confronting women and prosocial peer influence on their behaviors.

Moreover, the intervention increased communication about HIV/STI risk between the women and their partners. Although more than half of the women had never discussed such risk at baseline, four out of five women in the intervention group discussed HIV/STI risk at two months. The intervention group was more likely to have such communication than the control group throughout the eight-month follow-up period. The increase in dual-method contraceptive use could have been underpinned by frequent communication regarding HIV/STI risk with partners. Failure to practice the dual-method use was not mostly due to women’s inability, but their partner’s unwillingness to use condoms.^11^ Peipert et al. underscored the importance of education for male partners in either individual or couple-based intervention for promoting the dual-method use.^11^ However, reaching out to male partners may be more difficult compared to providing education to women visiting family planning clinics. In the current intervention, women received the handout used in the initial dual-method contraceptive counseling and were encouraged to discuss condom use with their partners. A qualitative study on married couples in Uganda found that women were more likely to initiate discussion and persuade their male partners to use condoms.^27^ In addition, the findings of our preliminary study reported that women could talk more comfortably about sensitive topics, such as condom use, by sharing information they received as a conversation starter.^16^ The majority of women in this study were willing to share the health messages and discuss HIV/STI risk with their partners. Given that women using HECs visit health facilities presumably more frequently than men, educating them on the dual-method use and encouraging them to share such messages with their partners are effective strategies.

Despite the increase in dual-method contraceptive use, no significant difference was observed in pregnancy occurrence between the intervention and control groups at each follow-up point. In this study, many women started the dual-method use but practiced it inconsistently. Inconsistent dual-method contraceptive use may explain the lack of effect on avoiding pregnancies.^28^ It might also be explained by a lack of statistical power. Only 21 women (about 2% of the participants) became pregnant during the eight-month follow-up. The low incidence of pregnancy is reasonable because we recruited women using an HEC and who wanted to avoid pregnancy at baseline. However, the intervention group showed the lower incidence of pregnancy over time. Thus, a further trial with a larger sample size is recommended to examine the effect of the intervention on the incidence of pregnancy.

The positive deviance approach can be effective in promoting dual-method contraceptive use among married or in-union women. Few intervention studies have demonstrated an increase in the dual-method use,^12,13^ and adherence to such practice was frequently low.^11^ Condom use is often considered awkward or unacceptable in long-term relationships, especially when women are using HECs.^8,10^ Moreover, condom use is typically considered a male responsibility.^29^ The positive deviance intervention can be effective in changing such norms. During counseling and workshop, the PDs shared their experiences to help participants realize that condom use is normal even among married or in-union women using HECs. Moreover, the intervention included role-playing exercises to enable women to negotiate condom use with their partners and instructions on correct condom use and sources of different types of condoms. The positive deviance intervention can empower women with the skills necessary for playing a proactive role in negotiation and condom use with their partners.

The study has several limitations. First, the study measured outcomes based on self-reports from the participants. Therefore, it is subject to measurement errors. Especially, dual-method contraceptive use could have been over-reported given the information provided to participants during the intervention. Women in the intervention group had longer contacts with the PDs, including a five-hour workshop, whereas those in the control group had only telephone-based contacts after the initial clinic-based counseling. Frequent contact in the intervention group may have resulted in the over-reporting of outcomes, which can lead to overestimating the intervention effect. Nevertheless, over-reporting of outcomes was minimized by assuring the participants of the confidentiality of their responses and conducting interviews by experienced female research assistants. Second, we collected data on pregnancy incidence during follow-up, but the rate was too low to use as a proxy for the dual-method use. Other clinical meaningful data, such as the incidence of STI, should be collected to evaluate interventions for the dual-method use in future research. Third, several characteristics of the participants were imbalanced between the intervention and control groups due to the relatively small number of clusters. However, random-effect model analysis was performed by controlling for cluster effects and differences in baseline characteristics to evaluate the effects of the intervention. Lastly, the intervention was developed based on the qualitative study of PDs in Mbarara District and examined its effectiveness among women in the same area. Just applying the intervention to other communities might not be effective. Each community may have different local solutions, group dynamics, and stakeholders.^30^ Therefore, they must participate in the process to identify their own solutions. Those solutions will be different from the ones identified in this study. Further research is recommended to assess the effectiveness of the positive deviance approach in a given context with careful attention to its process.^30^

## Conclusion

In summary, the positive deviance intervention was effective in promoting dual-method contraceptive use among married or in-union women using HECs in Mbarara District, Uganda. Condom use is often considered a male responsibility and unacceptable in long-term relationships, especially when women are using HECs. The positive deviance intervention can be a potential option for promoting the dual-method use by empowering women to play a proactive role in negotiation and condom use with their partners despite such social norms.

## Data Availability

The data underlying this study have been uploaded to the Figshare Repository and are accessible at https://doi.org/10.6084/m9.figshare.12936857.v1.

## Acknowledgements

The authors would like to thank all participants for their time and cooperation and extend their appreciation to the women joined in the trial as PDs and the research assistants.

## Footnotes

### Contributors

HK and MJ conceived the study and contributed to funding acquisition. HK, AS, JK, KICO, and MJ contributed to the study design. HK conducted the literature review. HK, CM, and SM led the development of the data collection instrument, data collection, and quality assessment. HK and AS did the statistical analysis. All authors contributed to data interpretation. HK wrote the original draft. AS, JK, KICO, SM, CM, and MJ reviewed and revised the manuscript. All authors approved the final version for submission.

### Patient consent

Not required.

### Ethics approval

The study was approved by the Research Ethics Committee of the Graduate School of Medicine, University of Tokyo (2019085NI), Institutional Research and Ethics Committee of Mbarara University of Science and Technology (IRB15/06-19), and Uganda National Council of Science and Technology (HS439ES).

### Provenance and peer review

Not commissioned; externally peer reviewed.

