## Supplementary Table S2 for "Positive deviance for promoting dual-method contraceptive use among women in Uganda: a cluster randomized controlled trial"

**Supplementary Table S2. Baseline characteristics of women lost to follow up in the intervention and control groups**

**Month 2**

| Variables | Intervention | | | | | Control | | | | |
|---|---|---|---|---|---|---|---|---|---|---|
| | Reached | | Lost to follow-up | | | Reached | | Lost to follow-up | | |
| | n | % | n | % | p-value† | n | % | n | % | p-value† |
| **1) Socio-demographic characteristics** | | | | | | | | | | |
| **Age in years, mean (SD)** | 30.6 | 6.5 | 29.8 | 6.7 | 0.291 | 30.5 | 5.8 | 27.7 | 5.8 | **<0.001** |
| **Education** | | | | | | | | | | |
| Never | 114 | 29.8 | 31 | 32.0 | 0.674 | 78 | 22.2 | 39 | 30.2 | 0.070 |
| Primary and more | 269 | 70.2 | 66 | 68.0 | | 273 | 77.8 | 90 | 69.8 | |
| **Religion** | | | | | | | | | | |
| Christian | 356 | 93.0 | 94 | 96.9 | 0.150 | 320 | 91.2 | 116 | 89.9 | 0.675 |
| Muslim | 27 | 7.1 | 3 | 3.1 | | 31 | 8.8 | 13 | 10.1 | |
| **Wealth index** | | | | | | | | | | |
| Poor | 139 | 36.29 | 37 | 38.1 | 0.821 | 119 | 33.9 | 39 | 30.2 | 0.070 |
| Middle | 133 | 34.73 | 35 | 36.1 | | 93 | 26.5 | 48 | 37.2 | |
| Rich | 111 | 28.98 | 25 | 25.8 | | 139 | 39.6 | 42 | 32.6 | |
| **No. of children, mean (SD)** | 3.2 | 1.7 | 3.0 | 1.7 | 0.428 | 3.0 | 1.9 | 2.7 | 1.7 | 0.099 |
| **Pregnancy intention** | | | | | | | | | | |
| No | 81 | 21.15 | 19 | 19.6 | 0.891 | 76 | 21.65 | 20 | 15.5 | 0.195 |
| Yes | 271 | 70.76 | 71 | 73.2 | | 247 | 70.37 | 94 | 72.9 | |
| Don't know | 31 | 8.09 | 7 | 7.2 | | 28 | 7.98 | 15 | 11.6 | |
| **Partner's pregnancy intention** | | | | | | | | | | |
| No | 57 | 14.88 | 12 | 12.4 | 0.462 | 53 | 15.1 | 15 | 11.6 | 0.293 |
| Yes | 259 | 67.62 | 63 | 65.0 | | 235 | 66.95 | 96 | 74.4 | |
| Don't know | 67 | 17.49 | 22 | 22.7 | | 63 | 17.95 | 18 | 14.0 | |
| **History of unintended pregnancy** | | | | | | | | | | |
| No | 245 | 63.97 | 68 | 70.1 | 0.257 | 248 | 70.66 | 87 | 67.4 | 0.497 |
| Yes | 138 | 36.03 | 29 | 29.9 | | 103 | 29.34 | 42 | 32.6 | |
| **Multiple sex partners** | | | | | | | | | | |
| No | 360 | 93.99 | 92 | 94.9 | 0.750 | 333 | 94.87 | 123 | 95.4 | 0.832 |
| Yes | 23 | 6.01 | 5 | 5.2 | | 18 | 5.13 | 6 | 4.7 | |
| **2) HIV-related characteristics** | | | | | | | | | | |
| **HIV status** | | | | | | | | | | |
| Negative | 351 | 91.64 | 87 | 89.7 | 0.543 | 123 | 89.17 | 313 | 95.4 | **0.038** |
| Positive | 32 | 8.36 | 10 | 10.3 | | 6 | 10.83 | 38 | 4.7 | |
| **Partner's HIV status** | | | | | | | | | | |
| Negative | 308 | 80.42 | 78 | 80.4 | 0.834 | 107 | 75.78 | 266 | 83.0 | 0.163 |
| Positive | 26 | 6.79 | 8 | 8.3 | | 6 | 9.4 | 33 | 4.7 | |
| Don't know | 49 | 12.79 | 11 | 11.3 | | 16 | 14.81 | 52 | 12.4 | |
| **Disclosure of HIV status** | | | | | | | | | | |
| No | 18 | 4.7 | 3 | 3.1 | 0.489 | 4 | 4.27 | 15 | 3.1 | 0.559 |
| Yes | 365 | 95.3 | 94 | 96.9 | | 125 | 95.73 | 336 | 96.9 | |
| **Perceived risk for HIV/STIs** | | | | | | | | | | |
| No risk at all | 47 | 12.27 | 15 | 15.5 | 0.748 | 20 | 19.09 | 67 | 15.5 | 0.789 |
| Small | 140 | 36.55 | 37 | 38.1 | | 48 | 37.04 | 130 | 37.2 | |
| Moderate | 112 | 29.24 | 24 | 24.7 | | 34 | 25.64 | 90 | 26.4 | |
| Great | 84 | 21.93 | 21 | 21.7 | | 27 | 18.23 | 64 | 20.9 | |
| **3) HEC use** | | | | | | | | | | |
| **Type of HECs** | | | | | | | | | | |
| Injectables | 198 | 51.7 | 54 | 55.7 | 0.901 | 178 | 50.71 | 68 | 52.7 | 0.910 |
| Implants | 127 | 33.16 | 28 | 28.9 | | 107 | 30.48 | 41 | 31.8 | |
| IUD | 34 | 8.88 | 9 | 9.3 | | 41 | 11.68 | 13 | 10.1 | |
| Pill | 22 | 5.74 | 5 | 5.2 | | 24 | 6.84 | 7 | 5.4 | |
| Female sterilization | 2 | 0.52 | 1 | 1.0 | | 1 | 0.28 | 0 | 0.0 | |
| **Partner's recognition of HEC use** | | | | | | | | | | |
| No | 32 | 8.36 | 4 | 4.1 | 0.158 | 29 | 8.26 | 14 | 10.9 | 0.378 |
| Yes | 351 | 91.64 | 93 | 95.9 | | 322 | 91.74 | 115 | 89.2 | |
| **Partner's attitude about HEC use** | | | | | | | | | | |
| Positive | 344 | 89.82 | 88 | 90.7 | 0.795 | 320 | 91.17 | 119 | 92.3 | 0.838 |
| Negative | 30 | 7.83 | 6 | 6.2 | | 26 | 7.41 | 9 | 7.0 | |
| Don't know | 9 | 2.35 | 3 | 3.1 | | 5 | 1.42 | 1 | 0.8 | |
| **4) Other psychosocial charactericts** | | | | | | | | | | |
| **HIV-related knowledge (HIV-KQ-18), mean (SD)** | 12.0 | 2.6 | 11.9 | 2.6 | 0.829 | 11.3 | 3.0 | 11.1 | 2.8 | 0.597 |
| **Condom use self-efficacy scale, mean (SD)** | 22.3 | 9.2 | 22.1 | 9.9 | 0.794 | 21.6 | 8.4 | 23.2 | 7.9 | 0.058 |
| **Sexual Relationship Power Scale** | | | | | | | | | | |
| Low | 141 | 36.81 | 32 | 33.0 | 0.484 | 114 | 32.48 | 38 | 29.5 | 0.763 |
| Medium | 129 | 33.68 | 39 | 40.2 | | 130 | 37.04 | 52 | 40.3 | |
| High | 113 | 29.5 | 26 | 26.8 | | 107 | 30.48 | 39 | 30.2 | |

**Month 4**

| Variables | Intervention | | | | | Control | | | | |
|---|---|---|---|---|---|---|---|---|---|---|
| | Reached | | Lost to follow-up | | | Reached | | Lost to follow-up | | |
| | n | % | n | % | p-value† | n | % | n | % | p-value† |
| **1) Socio-demographic characteristics** | | | | | | | | | | |
| **Age in years, mean (SD)** | 30.4 | 6.4 | 30.5 | 6.9 | 0.965 | 30.2 | 6.9 | 28.1 | 6.1 | **0.006** |
| **Education** | | | | | | | | | | |
| Never | 122 | 30.1 | 23 | 31.1 | 0.859 | 88 | 23.1 | 29 | 29.3 | 0.201 |
| Primary and more | 284 | 70.0 | 51 | 68.9 | | 293 | 76.9 | 70 | 70.7 | |
| **Religion** | | | | | | | | | | |
| Christian | 378 | 93.1 | 72 | 97.3 | 0.170 | 345 | 90.6 | 91 | 91.9 | 0.674 |
| Muslim | 28 | 6.9 | 2 | 2.7 | | 36 | 9.5 | 8 | 8.1 | |
| **Wealth index** | | | | | | | | | | |
| Poor | 142 | 35.0 | 34 | 46.0 | 0.190 | 133 | 34.9 | 25 | 25.3 | 0.188 |
| Middle | 145 | 35.7 | 23 | 31.1 | | 108 | 28.4 | 33 | 33.3 | |
| Rich | 119 | 29.3 | 17 | 23.0 | | 140 | 36.8 | 41 | 41.4 | |
| **No. of children, mean (SD)** | 3.2 | 1.7 | 3.1 | 1.9 | 0.859 | 3.0 | 1.9 | 2.6 | 1.7 | **0.044** |
| **Pregnancy intention** | | | | | | | | | | |
| No | 83 | 20.4 | 17 | 23.0 | 0.391 | 78 | 20.5 | 18 | 18.2 | 0.126 |
| Yes | 288 | 70.9 | 54 | 73.0 | | 274 | 71.9 | 67 | 67.7 | |
| Don't know | 35 | 8.6 | 3 | 4.1 | | 29 | 7.6 | 14 | 14.1 | |
| **Partner's pregnancy intention** | | | | | | | | | | |
| No | 55 | 13.6 | 14 | 18.9 | 0.454 | 57 | 15.0 | 11 | 11.1 | 0.541 |
| Yes | 276 | 68.0 | 46 | 62.2 | | 262 | 68.8 | 69 | 69.7 | |
| Don't know | 75 | 18.5 | 14 | 18.9 | | 62 | 16.3 | 19 | 19.2 | |
| **History of unintended pregnancy** | | | | | | | | | | |
| No | 266 | 65.5 | 47 | 63.5 | 0.739 | 265 | 69.6 | 70 | 70.7 | 0.824 |
| Yes | 140 | 34.5 | 27 | 36.5 | | 116 | 30.5 | 29 | 29.3 | |
| **Multiple sex partners** | | | | | | | | | | |
| No | 382 | 94.1 | 70 | 89.2 | 0.864 | 361 | 94.8 | 95 | 96.0 | 0.623 |
| Yes | 24 | 5.9 | 4 | 10.8 | | 20 | 5.3 | 4 | 4.0 | |
| **2) HIV-related characteristics** | | | | | | | | | | |
| **HIV status** | | | | | | | | | | |
| Negative | 372 | 91.6 | 66 | 89.7 | 0.495 | 343 | 90.0 | 93 | 93.9 | 0.229 |
| Positive | 34 | 8.4 | 8 | 10.3 | | 38 | 10.0 | 6 | 6.1 | |
| **Partner's HIV status** | | | | | | | | | | |
| Negative | 326 | 80.3 | 60 | 81.1 | 0.509 | 289 | 75.9 | 84 | 84.9 | 0.122 |
| Positive | 27 | 6.7 | 7 | 9.5 | | 35 | 9.2 | 4 | 4.0 | |
| Don't know | 53 | 13.1 | 7 | 9.5 | | 57 | 15.0 | 11 | 11.1 | |
| **Disclosure of HIV status** | | | | | | | | | | |
| No | 19 | 4.7 | 2 | 2.7 | 0.444 | 17 | 4.5 | 2 | 2.0 | 0.267 |
| Yes | 387 | 95.3 | 72 | 97.3 | | 364 | 95.5 | 97 | 98.0 | |
| **Perceived risk for HIV/STIs** | | | | | | | | | | |
| No risk at all | 47 | 11.6 | 15 | 20.3 | 0.193 | 71 | 18.6 | 16 | 16.2 | 0.922 |
| Small | 154 | 37.9 | 23 | 31.1 | | 142 | 37.3 | 36 | 36.4 | |
| Moderate | 117 | 28.8 | 19 | 25.7 | | 97 | 25.5 | 27 | 27.3 | |
| Great | 88 | 21.7 | 17 | 23.0 | | 71 | 18.6 | 20 | 20.2 | |
| **3) HEC use** | | | | | | | | | | |
| **Type of HECs** | | | | | | | | | | |
| Injectables | 207 | 51.0 | 45 | 60.8 | 0.478 | 190 | 49.9 | 56 | 56.6 | 0.673 |
| Implants | 135 | 33.3 | 20 | 27.0 | | 120 | 31.5 | 28 | 28.3 | |
| IUD | 38 | 9.4 | 5 | 6.8 | | 43 | 11.3 | 11 | 11.1 | |
| Pill | 24 | 5.9 | 3 | 4.1 | | 27 | 7.1 | 4 | 4.0 | |
| Female sterilization | 2 | 0.5 | 1 | 1.4 | | 1 | 0.3 | 0 | 0.0 | |
| **Partner's recognition of HEC use** | | | | | | | | | | |
| No | 34 | 8.4 | 2 | 2.7 | 0.088 | 33 | 8.7 | 10 | 10.1 | 0.655 |
| Yes | 372 | 91.6 | 72 | 97.3 | | 348 | 91.3 | 89 | 89.9 | |
| **Partner's attitude about HEC use** | | | | | | | | | | |
| Positive | 363 | 89.4 | 69 | 93.2 | 0.472 | 350 | 91.9 | 89 | 89.9 | 0.725 |
| Negative | 33 | 8.1 | 3 | 4.1 | | 26 | 6.8 | 9 | 9.1 | |
| Don't know | 10 | 2.5 | 2 | 2.7 | | 5 | 1.3 | 1 | 1.0 | |
| **4) Other psychosocial charactericts** | | | | | | | | | | |
| **HIV-related knowledge (HIV-KQ-18), mean (SD)** | 11.9 | 2.6 | 12.1 | 2.6 | 0.632 | 11.2 | 3.0 | 11.4 | 2.9 | 0.563 |
| **Condom use self-efficacy scale, mean (SD)** | 22.0 | 9.4 | 23.6 | 9.0 | 0.193 | 21.7 | 8.4 | 23.4 | 8.0 | 0.080 |
| **Sexual Relationship Power Scale** | | | | | | | | | | |
| Low | 151 | 37.2 | 22 | 29.7 | 0.416 | 123 | 32.3 | 29 | 29.3 | 0.146 |
| Medium | 138 | 34.0 | 30 | 40.5 | | 150 | 39.4 | 32 | 32.3 | |
| High | 117 | 28.8 | 22 | 29.7 | | 108 | 28.4 | 38 | 38.4 | |

**Month 6**

| Variables | Intervention | | | | | Control | | | | |
|---|---|---|---|---|---|---|---|---|---|---|
| | Reached | | Lost to follow-up | | | Reached | | Lost to follow-up | | |
| | n | % | n | % | p-value† | n | % | n | % | p-value† |
| **1) Socio-demographic characteristics** | | | | | | | | | | |
| **Age in years, mean (SD)** | 30.6 | 6.4 | 29.5 | 6.8 | 0.166 | 30.1 | 6.9 | 28.3 | 6.2 | **0.016** |
| **Education** | | | | | | | | | | |
| Never | 118 | 30.0 | 27 | 31.4 | 0.791 | 88 | 22.9 | 29 | 30.5 | 0.119 |
| Primary and more | 276 | 70.1 | 59 | 68.6 | | 297 | 77.1 | 66 | 69.5 | |
| **Religion** | | | | | | | | | | |
| Christian | 366 | 92.9 | 84 | 96.9 | 0.097 | 349 | 90.7 | 87 | 91.6 | 0.779 |
| Muslim | 28 | 7.1 | 2 | 3.1 | | 36 | 9.4 | 8 | 8.4 | |
| **Wealth index** | | | | | | | | | | |
| Poor | 139 | 35.3 | 37 | 43.0 | 0.134 | 135 | 35.1 | 23 | 24.2 | 0.119 |
| Middle | 136 | 34.5 | 32 | 37.2 | | 111 | 28.8 | 30 | 31.6 | |
| Rich | 119 | 30.2 | 17 | 19.8 | | 139 | 36.1 | 42 | 44.2 | |
| **No. of children, mean (SD)** | 3.2 | 1.7 | 3.0 | 1.8 | 0.394 | 3.0 | 1.8 | 2.8 | 1.9 | 0.343 |
| **Pregnancy intention** | | | | | | | | | | |
| No | 84 | 21.3 | 16 | 18.6 | 0.349 | 77 | 20.0 | 19 | 20.0 | 0.190 |
| Yes | 276 | 70.1 | 66 | 76.7 | | 278 | 72.2 | 63 | 66.3 | |
| Don't know | 34 | 8.6 | 4 | 4.7 | | 30 | 7.8 | 13 | 13.7 | |
| **Partner's pregnancy intention** | | | | | | | | | | |
| No | 58 | 14.7 | 11 | 12.8 | 0.896 | 55 | 14.3 | 13 | 13.7 | 0.833 |
| Yes | 263 | 66.8 | 59 | 68.6 | | 267 | 69.4 | 64 | 67.4 | |
| Don't know | 73 | 18.5 | 16 | 18.6 | | 63 | 16.4 | 18 | 19.0 | |
| **History of unintended pregnancy** | | | | | | | | | | |
| No | 258 | 65.5 | 55 | 64.0 | 0.787 | 265 | 68.8 | 70 | 73.7 | 0.356 |
| Yes | 136 | 34.5 | 31 | 36.1 | | 120 | 31.2 | 25 | 26.3 | |
| **Multiple sex partners** | | | | | | | | | | |
| No | 371 | 94.2 | 81 | 94.2 | 0.993 | 365 | 94.8 | 91 | 95.8 | 0.693 |
| Yes | 23 | 5.8 | 5 | 5.8 | | 20 | 5.2 | 4 | 4.2 | |
| **2) HIV-related characteristics** | | | | | | | | | | |
| **HIV status** | | | | | | | | | | |
| Negative | 359 | 91.1 | 79 | 91.9 | 0.825 | 347 | 90.1 | 89 | 93.7 | 0.282 |
| Positive | 35 | 8.9 | 7 | 8.1 | | 38 | 9.9 | 6 | 6.3 | |
| **Partner's HIV status** | | | | | | | | | | |
| Negative | 313 | 79.4 | 73 | 84.9 | 0.225 | 295 | 76.6 | 78 | 82.1 | 0.431 |
| Positive | 27 | 6.9 | 7 | 8.1 | | 34 | 8.8 | 5 | 5.3 | |
| Don't know | 54 | 13.7 | 6 | 7.0 | | 56 | 14.6 | 12 | 12.6 | |
| **Disclosure of HIV status** | | | | | | | | | | |
| No | 18 | 4.6 | 3 | 3.5 | 0.657 | 16 | 4.2 | 3 | 3.2 | 0.655 |
| Yes | 376 | 95.4 | 83 | 96.5 | | 369 | 95.8 | 92 | 96.8 | |
| **Perceived risk for HIV/STIs** | | | | | | | | | | |
| No risk at all | 43 | 10.9 | 19 | 22.1 | **0.044** | 70 | 18.2 | 17 | 17.9 | 0.949 |
| Small | 148 | 37.6 | 29 | 33.7 | | 145 | 37.7 | 33 | 34.7 | |
| Moderate | 116 | 29.4 | 20 | 23.3 | | 98 | 25.5 | 26 | 27.4 | |
| Great | 87 | 22.1 | 18 | 20.9 | | 72 | 18.7 | 19 | 20.0 | |
| **3) HEC use** | | | | | | | | | | |
| **Type of HECs** | | | | | | | | | | |
| Injectables | 193 | 49.0 | 59 | 68.6 | **0.010** | 190 | 49.4 | 56 | 59.0 | 0.380 |
| Implants | 135 | 34.3 | 20 | 23.3 | | 120 | 31.2 | 28 | 29.5 | |
| IUD | 38 | 9.6 | 5 | 5.8 | | 47 | 12.2 | 7 | 7.4 | |
| Pill | 26 | 6.6 | 1 | 1.2 | | 27 | 7.0 | 4 | 4.2 | |
| Female sterilization | 2 | 0.5 | 1 | 1.2 | | 1 | 0.3 | 0 | 0.0 | |
| **Partner's recognition of HEC use** | | | | | | | | | | |
| No | 34 | 8.6 | 2 | 2.3 | **0.044** | 32 | 8.3 | 11 | 11.6 | 0.318 |
| Yes | 360 | 91.4 | 84 | 97.7 | | 353 | 91.7 | 84 | 88.4 | |
| **Partner's attitude about HEC use** | | | | | | | | | | |
| Positive | 351 | 89.1 | 81 | 94.2 | 0.292 | 353 | 91.7 | 86 | 90.5 | 0.880 |
| Negative | 33 | 8.4 | 3 | 3.5 | | 27 | 7.0 | 8 | 8.4 | |
| Don't know | 10 | 2.5 | 2 | 2.3 | | 5 | 1.3 | 1 | 1.1 | |
| **4) Other psychosocial charactericts** | | | | | | | | | | |
| **HIV-related knowledge (HIV-KQ-18), mean (SD)** | 12.0 | 2.6 | 11.8 | 2.8 | 0.507 | 11.3 | 3.0 | 11.2 | 2.9 | 0.946 |
| **Condom use self-efficacy scale, mean (SD)** | 22.1 | 9.3 | 22.9 | 9.7 | 0.478 | 21.9 | 8.2 | 22.7 | 8.7 | 0.422 |
| **Sexual Relationship Power Scale** | | | | | | | | | | |
| Low | 144 | 36.6 | 29 | 33.7 | 0.561 | 121 | 31.4 | 31 | 29.5 | 0.442 |
| Medium | 140 | 35.5 | 28 | 32.6 | | 151 | 39.2 | 31 | 40.3 | |
| High | 110 | 27.9 | 29 | 33.7 | | 113 | 29.4 | 33 | 30.2 | |

**Month 8**

| Variables | Intervention | | | | | Control | | | | |
|---|---|---|---|---|---|---|---|---|---|---|
| | Reached | | Lost to follow-up | | | Reached | | Lost to follow-up | | |
| | n | % | n | % | p-value† | n | % | n | % | p-value† |
| **1) Socio-demographic characteristics** | | | | | | | | | | |
| **Age in years, mean (SD)** | 30.6 | 6.4 | 29.5 | 7.2 | 0.158 | 30.1 | 6.9 | 28.5 | 6.1 | **0.044** |
| **Education** | | | | | | | | | | |
| Never | 119 | 29.4 | 26 | 34.7 | 0.360 | 87 | 22.6 | 30 | 31.6 | 0.068 |
| Primary and more | 286 | 70.6 | 49 | 65.3 | | 298 | 77.4 | 65 | 68.4 | |
| **Religion** | | | | | | | | | | |
| Christian | 376 | 92.8 | 74 | 98.7 | 0.055 | 349 | 90.7 | 87 | 91.6 | 0.779 |
| Muslim | 29 | 7.2 | 1 | 1.3 | | 36 | 9.4 | 8 | 8.4 | |
| **Wealth index** | | | | | | | | | | |
| Poor | 143 | 35.3 | 33 | 44.0 | 0.112 | 135 | 35.1 | 23 | 24.2 | 0.119 |
| Middle | 140 | 34.6 | 28 | 37.3 | | 111 | 28.8 | 30 | 31.6 | |
| Rich | 122 | 30.1 | 14 | 18.7 | | 139 | 36.1 | 42 | 44.2 | |
| **No. of children, mean (SD)** | 3.2 | 1.7 | 3.0 | 1.9 | 0.438 | 2.9 | 1.9 | 2.8 | 1.8 | 0.607 |
| **Pregnancy intention** | | | | | | | | | | |
| No | 88 | 21.7 | 12 | 16.0 | 0.442 | 21 | 22.1 | 19.48 | 22.1 | 0.462 |
| Yes | 284 | 70.1 | 58 | 77.3 | | 63 | 66.3 | 72.21 | 66.3 | |
| Don't know | 33 | 8.2 | 5 | 6.7 | | 11 | 11.6 | 8.31 | 11.6 | |
| **Partner's pregnancy intention** | | | | | | | | | | |
| No | 61 | 15.1 | 8 | 10.7 | 0.600 | 14 | 14.7 | 14.03 | 14.7 | 0.799 |
| Yes | 269 | 66.4 | 53 | 70.7 | | 63 | 66.3 | 69.61 | 66.3 | |
| Don't know | 75 | 18.5 | 14 | 18.7 | | 18 | 19.0 | 16.36 | 19.0 | |
| **History of unintended pregnancy** | | | | | | | | | | |
| No | 261 | 64.4 | 52 | 69.3 | 0.414 | 69 | 72.6 | 69.09 | 72.6 | 0.501 |
| Yes | 144 | 35.6 | 23 | 30.7 | | 26 | 27.4 | 30.91 | 27.4 | |
| **Multiple sex partners** | | | | | | | | | | |
| No | 380 | 93.8 | 72 | 96.0 | 0.461 | 91 | 95.8 | 94.81 | 95.8 | 0.693 |
| Yes | 25 | 6.2 | 3 | 4.0 | | 4 | 4.2 | 5.19 | 4.2 | |
| **2) HIV-related characteristics** | | | | | | | | | | |
| **HIV status** | | | | | | | | | | |
| Negative | 372 | 91.9 | 66 | 88.0 | 0.278 | 347 | 90.1 | 89 | 93.7 | 0.282 |
| Positive | 33 | 8.2 | 9 | 12.0 | | 38 | 9.9 | 6 | 6.3 | |
| **Partner's HIV status** | | | | | | | | | | |
| Negative | 324 | 80.0 | 62 | 82.7 | 0.646 | 295 | 76.6 | 78 | 82.1 | 0.431 |
| Positive | 28 | 6.9 | 6 | 8.0 | | 34 | 8.8 | 5 | 5.3 | |
| Don't know | 53 | 13.1 | 7 | 9.3 | | 56 | 14.6 | 12 | 12.6 | |
| **Disclosure of HIV status** | | | | | | | | | | |
| No | 20 | 4.9 | 1 | 1.3 | 0.161 | 15 | 3.9 | 4 | 4.2 | 0.888 |
| Yes | 385 | 95.1 | 74 | 98.7 | | 370 | 96.1 | 91 | 95.8 | |
| **Perceived risk for HIV/STIs** | | | | | | | | | | |
| No risk at all | 48 | 11.9 | 14 | 18.7 | 0.395 | 70 | 18.2 | 17 | 17.9 | 0.999 |
| Small | 149 | 36.8 | 28 | 37.3 | | 143 | 37.1 | 35 | 36.8 | |
| Moderate | 117 | 28.9 | 19 | 25.3 | | 99 | 25.7 | 25 | 26.3 | |
| Great | 91 | 22.5 | 14 | 18.7 | | 73 | 19.0 | 18 | 19.0 | |
| **3) HEC use** | | | | | | | | | | |
| **Type of HECs** | | | | | | | | | | |
| Injectables | 204 | 50.4 | 48 | 64.0 | 0.199 | 190 | 49.4 | 56 | 59.0 | 0.450 |
| Implants | 136 | 33.6 | 19 | 25.3 | | 121 | 31.4 | 27 | 28.4 | |
| IUD | 39 | 9.6 | 4 | 5.3 | | 47 | 12.2 | 7 | 7.4 | |
| Pill | 24 | 5.9 | 3 | 4.0 | | 26 | 6.8 | 5 | 5.3 | |
| Female sterilization | 2 | 0.5 | 1 | 1.3 | | 1 | 0.3 | 0 | 0.0 | |
| **Partner's recognition of HEC use** | | | | | | | | | | |
| No | 35 | 8.6 | 1 | 1.3 | **0.027** | 32 | 8.3 | 11 | 11.6 | 0.318 |
| Yes | 370 | 91.4 | 74 | 98.7 | | 353 | 91.7 | 84 | 88.4 | |
| **Partner's attitude about HEC use** | | | | | | | | | | |
| Positive | 361 | 89.1 | 71 | 94.7 | 0.224 | 353 | 91.7 | 86 | 90.5 | 0.880 |
| Negative | 34 | 8.4 | 2 | 2.7 | | 27 | 7.0 | 8 | 8.4 | |
| Don't know | 10 | 2.5 | 2 | 2.7 | | 5 | 1.3 | 1 | 1.1 | |
| **4) Other psychosocial charactericts** | | | | | | | | | | |
| **HIV-related knowledge (HIV-KQ-18), mean (SD)** | 12.0 | 2.6 | 11.8 | 2.4 | 0.527 | 11.2 | 3.0 | 11.3 | 2.8 | 0.992 |
| **Condom use self-efficacy scale, mean (SD)** | 22.3 | 9.2 | 22.1 | 10.2 | 0.848 | 22.0 | 8.2 | 22.4 | 8.8 | 0.608 |
| **Sexual Relationship Power Scale** | | | | | | | | | | |
| Low | 146 | 36.1 | 27 | 36.0 | 0.327 | 122 | 31.7 | 30 | 31.6 | 0.526 |
| Medium | 137 | 33.8 | 31 | 41.3 | | 150 | 39.0 | 32 | 33.7 | |
| High | 122 | 30.1 | 17 | 22.7 | | 113 | 29.4 | 33 | 34.7 | |

SD: standard deviation; HIV: human immunodeficiency virus; STI: sexually transmitted infection; HEC: highly effective contraceptive; IUD: intrauterine device

† Based on Chi-squared test for categorical variables and t-test for continuous variables.
