## Supplementary Tables S3-S17 for "Positive deviance for promoting dual-method contraceptive use among women in Uganda: a cluster randomized controlled trial"

**Supplementary Table S3. Effects of intervention on dual-method use at last sexual intercourse amonng women at 2 months after enrollement**

| Variables | Model 1 | | | Model 2 | | | Model 3 | | |
|---|---|---|---|---|---|---|---|---|---|
| | OR | (95% CI) | p-value | AOR[a] | (95% CI) | p-value | AOR[b] | (95% CI) | p-value |
| **Intervention** | | | | | | | | | |
| Control | Ref. | | | Ref. | | | Ref. | | |
| Intervention | 4.62 | ( 3.18 - 6.7125 ) | **<0.001** | 1.17 | ( 0.46 - 2.9811 ) | 0.745 | 1.19 | ( 0.48 - 2.95 ) | 0.712 |
| **Time** | | | | 2.76 | ( 1.63 - 4.6676 ) | **<0.001** | 2.89 | ( 1.70 - 4.89 ) | 1.712 |
| **Intervention*time[c]** | | | | 4.29 | ( 2.12 - 8.69 ) | **<0.001** | 4.12 | ( 2.02 - 8.39 ) | **<0.001** |
| **1) Socio-demographic characteristics** | | | | | | | | | |
| **Age in years** | | | | | | | 1.00 | ( 0.96 - 1.04 ) | 0.712 |
| **Education** | | | | | | | | | |
| Never | | | | | | | Ref. | | |
| Primary and more | | | | | | | 0.98 | ( 0.66 - 1.47 ) | 0.935 |
| **Religion** | | | | | | | | | |
| Christian | | | | | | | Ref. | | |
| Muslim | | | | | | | 1.42 | ( 0.78 - 2.58 ) | 0.246 |
| **Wealth index** | | | | | | | | | |
| Poor | | | | | | | Ref. | | |
| Middle | | | | | | | 1.35 | ( 0.89 - 2.05 ) | 0.164 |
| Rich | | | | | | | 1.31 | ( 0.84 - 2.05 ) | 0.24 |
| **No. of children** | | | | | | | 0.87 | ( 0.75 - 1.00 ) | 0.057 |
| **Pregnancy intention** | | | | | | | | | |
| No | | | | | | | Ref. | | |
| Yes | | | | | | | 1.17 | ( 0.66 - 2.09 ) | 0.592 |
| Don't know | | | | | | | 1.54 | ( 0.71 - 3.34 ) | 0.274 |
| **Partner's pregnancy intention** | | | | | | | | | |
| No | | | | | | | Ref. | | |
| Yes | | | | | | | 0.45 | ( 0.24 - 0.85 ) | **0.013** |
| Don't know | | | | | | | 0.49 | ( 0.25 - 0.96 ) | **0.038** |
| **History of unintended pregnancy** | | | | | | | | | |
| No | | | | | | | Ref. | | |
| Yes | | | | | | | 0.93 | ( 0.64 - 1.34 ) | 0.68 |
| **Multiple sex partners** | | | | | | | | | |
| No | | | | | | | Ref. | | |
| Yes | | | | | | | 3.50 | ( 1.85 - 6.62 ) | **<0.001** |
| **2) HIV-related characteristics** | | | | | | | | | |
| **HIV status** | | | | | | | | | |
| Negative | | | | | | | Ref. | | |
| Positive | | | | | | | 1.57 | ( 0.71 - 3.49 ) | 0.267 |
| **Partner's HIV status** | | | | | | | | | |
| Negative | | | | | | | Ref. | | |
| Positive | | | | | | | 1.27 | ( 0.54 - 2.99 ) | 0.583 |
| Don't know | | | | | | | 0.95 | ( 0.57 - 1.58 ) | 0.837 |
| **Perceived risk for HIV/STIs** | | | | | | | | | |
| No risk at all | | | | | | | Ref. | | |
| Small | | | | | | | 0.80 | ( 0.47 - 1.37 ) | 0.421 |
| Moderate | | | | | | | 1.05 | ( 0.60 - 1.83 ) | 0.858 |
| Great | | | | | | | 1.18 | ( 0.65 - 2.15 ) | 0.588 |
| **3) HEC use** | | | | | | | | | |
| **Type of HECs** | | | | | | | | | |
| Injectables | | | | | | | Ref. | | |
| Implants | | | | | | | 0.94 | ( 0.65 - 1.35 ) | 0.726 |
| IUD | | | | | | | 1.21 | ( 0.69 - 2.12 ) | 0.505 |
| Pill | | | | | | | 0.83 | ( 0.40 - 1.72 ) | 0.611 |
| Female sterilization | | | | | | | | | perfect success |
| **4) Other psychosocial charactericts** | | | | | | | | | |
| **HIV-related knowledge (HIV-KQ-18)** | | | | | | | 1.03 | ( 0.97 - 1.11 ) | 0.338 |
| **Condom use self-efficacy scale** | | | | | | | 1.02 | ( 1.00 - 1.05 ) | **0.035** |
| **Sexual Relationship Power Scale** | | | | | | | | | |
| Low | | | | | | | Ref. | | |
| Medium | | | | | | | 1.13 | ( 0.76 - 1.69 ) | 0.551 |
| High | | | | | | | 1.07 | ( 0.70 - 1.66 ) | 0.748 |

OR: odds ratio; AOR: adjusted odds ratio; SD: standard deviation; HIV: human immunodeficiency virus; STI: sexually transmitted infection; HEC: highly effective contraceptive; IUD: intrauterine device

a. Adjusted for the cluster effect

b. Adjusted for age, education, religion, wealth index, number of children, pregnancy intention, partner's pregnancy intention, history of unintended pregnancy, multiple sex partnership, HIV status, partner's HIV status, risk perception of HIV/STIs, HIV-related knowledge, condom use self-efficacy, and sexual relationship control power.

c. Intervention*time represents the status of the intervention group at follow-up in comparison with the control group at the baseline.

**Supplementary Table S4. Effects of intervention on dual-method use at last sexual intercourse amonng women at 4 months after enrollement**

| Variables | Model 1 | | | Model 2 | | | Model 3 | | |
|---|---|---|---|---|---|---|---|---|---|
| | OR | (95% CI) | p-value | AOR[a] | (95% CI) | p-value | AOR[b] | (95% CI) | p-value |
| **Intervention** | | | | | | | | | |
| Control | Ref. | | | Ref. | | | Ref. | | |
| Intervention | 2.13 | ( 1.49 - 3.0643 ) | **<0.001** | 1.62 | ( 0.81 - 3.2637 ) | 0.176 | 1.66 | ( 0.87 - 3.16 ) | 0.121 |
| **Time** | | | | 3.55 | ( 2.07 - 6.0778 ) | **<0.001** | 3.55 | ( 2.08 - 6.08 ) | **<0.001** |
| **Intervention*time[c]** | | | | 1.66 | ( 0.84 - 3.30 ) | 0.148 | 1.66 | ( 0.84 - 3.30 ) | 0.146 |
| **1) Socio-demographic characteristics** | | | | | | | | | |
| **Age in years** | | | | | | | 0.99 | ( 0.95 - 1.03 ) | 0.530 |
| **Education** | | | | | | | | | |
| Never | | | | | | | Ref. | | |
| Primary and more | | | | | | | 0.79 | ( 0.51 - 1.22 ) | 0.278 |
| **Religion** | | | | | | | | | |
| Christian | | | | | | | Ref. | | |
| Muslim | | | | | | | 1.28 | ( 0.66 - 2.49 ) | 0.465 |
| **Wealth index** | | | | | | | | | |
| Poor | | | | | | | Ref. | | |
| Middle | | | | | | | 1.12 | ( 0.71 - 1.76 ) | 0.624 |
| Rich | | | | | | | 1.14 | ( 0.70 - 1.85 ) | 0.608 |
| **No. of children** | | | | | | | 0.92 | ( 0.78 - 1.08 ) | 0.314 |
| **Pregnancy intention** | | | | | | | | | |
| No | | | | | | | Ref. | | |
| Yes | | | | | | | 0.75 | ( 0.40 - 1.42 ) | 0.376 |
| Don't know | | | | | | | 1.17 | ( 0.51 - 2.66 ) | 0.715 |
| **Partner's pregnancy intention** | | | | | | | | | |
| No | | | | | | | Ref. | | |
| Yes | | | | | | | 0.55 | ( 0.28 - 1.09 ) | 0.085 |
| Don't know | | | | | | | 0.55 | ( 0.26 - 1.15 ) | 0.113 |
| **History of unintended pregnancy** | | | | | | | | | |
| No | | | | | | | Ref. | | |
| Yes | | | | | | | 0.62 | ( 0.40 - 0.94 ) | 0.026 |
| **Multiple sex partners** | | | | | | | | | |
| No | | | | | | | Ref. | | |
| Yes | | | | | | | 2.87 | ( 1.45 - 5.67 ) | **0.002** |
| **2) HIV-related characteristics** | | | | | | | | | |
| **HIV status** | | | | | | | | | |
| Negative | | | | | | | Ref. | | |
| Positive | | | | | | | 1.61 | ( 0.69 - 3.80 ) | 0.273 |
| **Partner's HIV status** | | | | | | | | | |
| Negative | | | | | | | Ref. | | |
| Positive | | | | | | | 1.40 | ( 0.55 - 3.52 ) | 0.48 |
| Don't know | | | | | | | 1.26 | ( 0.74 - 2.15 ) | 0.389 |
| **Perceived risk for HIV/STIs** | | | | | | | | | |
| No risk at all | | | | | | | Ref. | | |
| Small | | | | | | | 0.84 | ( 0.47 - 1.49 ) | 0.544 |
| Moderate | | | | | | | 1.01 | ( 0.56 - 1.83 ) | 0.975 |
| Great | | | | | | | 0.96 | ( 0.50 - 1.84 ) | 0.894 |
| **3) HEC use** | | | | | | | | | |
| **Type of HECs** | | | | | | | | | |
| Injectables | | | | | | | Ref. | | |
| Implants | | | | | | | 0.94 | ( 0.62 - 1.44 ) | 0.788 |
| IUD | | | | | | | 1.18 | ( 0.63 - 2.21 ) | 0.603 |
| Pill | | | | | | | 2.35 | ( 1.17 - 4.74 ) | 0.017 |
| Female sterilization | | | | | | | 0.974144 | ( 0.05 - 19.29 ) | 0.99 |
| **4) Other psychosocial charactericts** | | | | | | | | | |
| **HIV-related knowledge (HIV-KQ-18)** | | | | | | | 1.01 | ( 0.94 - 1.08 ) | 0.858 |
| **Condom use self-efficacy scale** | | | | | | | 1.04 | ( 1.01 - 1.06 ) | **0.002** |
| **Sexual Relationship Power Scale** | | | | | | | | | |
| Low | | | | | | | Ref. | | |
| Medium | | | | | | | 1.44 | ( 0.91 - 2.27 ) | 0.119 |
| High | | | | | | | 1.21 | ( 0.74 - 1.98 ) | 0.443 |

OR: odds ratio; AOR: adjusted odds ratio; SD: standard deviation; HIV: human immunodeficiency virus; STI: sexually transmitted infection; HEC: highly effective contraceptive; IUD: intrauterine device

a. Adjusted for the cluster effect

b. Adjusted for age, education, religion, wealth index, number of children, pregnancy intention, partner's pregnancy intention, history of unintended pregnancy, multiple sex partnership, HIV status, partner's HIV status, risk perception of HIV/STIs, HIV-related knowledge, condom use self-efficacy, and sexual relationship control power.

c. Intervention*time represents the status of the intervention group at follow-up in comparison with the control group at the baseline.

**Supplementary Table S5. Effects of intervention on dual-method use at last sexual intercourse amonng women at 6 months after enrollement**

| Variables | Model 1 | | | Model 2 | | | Model 3 | | |
|---|---|---|---|---|---|---|---|---|---|
| | OR | (95% CI) | p-value | AOR[a] | (95% CI) | p-value | AOR[b] | (95% CI) | p-value |
| **Intervention** | | | | | | | | | |
| Control | Ref. | | | Ref. | | | Ref. | | |
| Intervention | 2.53 | ( 1.69 - 3.7922 ) | **<0.001** | 1.42 | ( 0.55 - 3.6676 ) | 0.465 | 1.40 | ( 0.53 - 3.67 ) | 0.494 |
| **Time** | | | | 2.16 | ( 1.24 - 3.7523 ) | **0.006** | 2.17 | ( 1.25 - 3.76 ) | **0.006** |
| **Intervention*time[c]** | | | | 2.04 | ( 1.00 - 4.17 ) | 0.051 | 2.03 | ( 0.99 - 4.14 ) | 0.052 |
| **1) Socio-demographic characteristics** | | | | | | | | | |
| **Age in years** | | | | | | | 0.97 | ( 0.93 - 1.02 ) | 0.208 |
| **Education** | | | | | | | | | |
| Never | | | | | | | Ref. | | |
| Primary and more | | | | | | | 0.89 | ( 0.56 - 1.41 ) | 0.618 |
| **Religion** | | | | | | | | | |
| Christian | | | | | | | Ref. | | |
| Muslim | | | | | | | 1.36 | ( 0.70 - 2.65 ) | 0.366 |
| **Wealth index** | | | | | | | | | |
| Poor | | | | | | | Ref. | | |
| Middle | | | | | | | 0.96 | ( 0.60 - 1.55 ) | 0.875 |
| Rich | | | | | | | 0.75 | ( 0.45 - 1.27 ) | 0.283 |
| **No. of children** | | | | | | | 1.02 | ( 0.86 - 1.20 ) | 0.853 |
| **Pregnancy intention** | | | | | | | | | |
| No | | | | | | | Ref. | | |
| Yes | | | | | | | 0.84 | ( 0.44 - 1.61 ) | 0.602 |
| Don't know | | | | | | | 1.29 | ( 0.55 - 3.03 ) | 0.565 |
| **Partner's pregnancy intention** | | | | | | | | | |
| No | | | | | | | Ref. | | |
| Yes | | | | | | | 0.69 | ( 0.34 - 1.41 ) | 0.307 |
| Don't know | | | | | | | 0.62 | ( 0.29 - 1.35 ) | 0.228 |
| **History of unintended pregnancy** | | | | | | | | | |
| No | | | | | | | Ref. | | |
| Yes | | | | | | | 0.73 | ( 0.48 - 1.13 ) | 0.157 |
| **Multiple sex partners** | | | | | | | | | |
| No | | | | | | | Ref. | | |
| Yes | | | | | | | 2.96 | ( 1.50 - 5.85 ) | **0.002** |
| **2) HIV-related characteristics** | | | | | | | | | |
| **HIV status** | | | | | | | | | |
| Negative | | | | | | | Ref. | | |
| Positive | | | | | | | 1.24 | ( 0.52 - 2.97 ) | 0.629 |
| **Partner's HIV status** | | | | | | | | | |
| Negative | | | | | | | Ref. | | |
| Positive | | | | | | | 1.67 | ( 0.64 - 4.31 ) | 0.292 |
| Don't know | | | | | | | 1.09 | ( 0.62 - 1.92 ) | 0.758 |
| **Perceived risk for HIV/STIs** | | | | | | | | | |
| No risk at all | | | | | | | Ref. | | |
| Small | | | | | | | 0.76 | ( 0.41 - 1.40 ) | 0.377 |
| Moderate | | | | | | | 1.03 | ( 0.55 - 1.92 ) | 0.937 |
| Great | | | | | | | 0.77 | ( 0.38 - 1.53 ) | 0.452 |
| **3) HEC use** | | | | | | | | | |
| **Type of HECs** | | | | | | | | | |
| Injectables | | | | | | | Ref. | | |
| Implants | | | | | | | 0.92 | ( 0.60 - 1.42 ) | 0.715 |
| IUD | | | | | | | 1.19 | ( 0.63 - 2.25 ) | 0.589 |
| Pill | | | | | | | 2.03 | ( 0.99 - 4.17 ) | 0.054 |
| Female sterilization | | | | | | | | | perfect success |
| **4) Other psychosocial characterícts** | | | | | | | | | |
| **HIV-related knowledge (HIV-KQ-18)** | | | | | | | 1.05 | ( 0.97 - 1.13 ) | 0.226 |
| **Condom use self-efficacy scale** | | | | | | | 1.04 | ( 1.01 - 1.06 ) | **0.006** |
| **Sexual Relationship Power Scale** | | | | | | | | | |
| Low | | | | | | | Ref. | | |
| Medium | | | | | | | 1.59 | ( 0.99 - 2.54 ) | 0.056 |
| High | | | | | | | 1.19 | ( 0.71 - 1.97 ) | 0.513 |

OR: odds ratio; AOR: adjusted odds ratio; SD: standard deviation; HIV: human immunodeficiency virus; STI: sexually transmitted infection; HEC: highly effective contraceptive; IUD: intrauterine device

a. Adjusted for the cluster effect

b. Adjusted for age, education, religion, wealth index, number of children, pregnancy intention, partner's pregnancy intention, history of unintended pregnancy, multiple sex partnership, HIV status, partner's HIV status, risk perception of HIV/STIs, HIV-related knowledge, condom use self-efficacy, and sexual relationship control power.

c. Intervention*time represents the status of the intervention group at follow-up in comparison with the control group at the baseline.

**Supplementary Table S6. Effects of intervention on dual-method use at last sexual intercourse amonng women at 8 months after enrollement**

| Variables | Model 1 | | | Model 2 | | | Model 3 | | |
|---|---|---|---|---|---|---|---|---|---|
| | OR | (95% CI) | p-value | AOR[a] | (95% CI) | p-value | AOR[b] | (95% CI) | p-value |
| **Intervention** | | | | | | | | | |
| Control | Ref. | | | Ref. | | | Ref. | | |
| Intervention | 2.76 | ( 1.79 - 4.2587 ) | **<0.001** | 1.41 | ( 0.58 - 3.40 ) | 0.450 | 1.39 | ( 0.59 - 3.31 ) | 0.452 |
| **Time** | | | | 1.59 | ( 0.91 - 2.76 ) | 0.101 | 1.60 | ( 0.92 - 2.77 ) | 0.094 |
| **Intervention*time[c]** | | | | 2.19 | ( 1.07 - 4.48 ) | **0.032** | 2.16 | ( 1.06 - 4.41 ) | **0.034** |
| **1) Socio-demographic characteristics** | | | | | | | | | |
| **Age in years** | | | | | | | 0.97 | ( 0.93 - 1.01 ) | 0.114 |
| **Education** | | | | | | | | | |
| Never | | | | | | | Ref. | | |
| Primary and more | | | | | | | 1.03 | ( 0.66 - 1.62 ) | 0.884 |
| **Religion** | | | | | | | | | |
| Christian | | | | | | | Ref. | | |
| Muslim | | | | | | | 1.13 | ( 0.57 - 2.21 ) | 0.728 |
| **Wealth index** | | | | | | | | | |
| Poor | | | | | | | Ref. | | |
| Middle | | | | | | | 1.11 | ( 0.71 - 1.75 ) | 0.647 |
| Rich | | | | | | | 0.89 | ( 0.54 - 1.48 ) | 0.664 |
| **No. of children** | | | | | | | 1.04 | ( 0.88 - 1.22 ) | 0.676 |
| **Pregnancy intention** | | | | | | | | | |
| No | | | | | | | Ref. | | |
| Yes | | | | | | | 0.82 | ( 0.44 - 1.55 ) | 0.550 |
| Don't know | | | | | | | 1.66 | ( 0.75 - 3.65 ) | 0.210 |
| **Partner's pregnancy intention** | | | | | | | | | |
| No | | | | | | | Ref. | | |
| Yes | | | | | | | 0.65 | ( 0.33 - 1.28 ) | 0.214 |
| Don't know | | | | | | | 0.71 | ( 0.35 - 1.47 ) | 0.359 |
| **History of unintended pregnancy** | | | | | | | | | |
| No | | | | | | | Ref. | | |
| Yes | | | | | | | 0.83 | ( 0.55 - 1.25 ) | 0.375 |
| **Multiple sex partners** | | | | | | | | | |
| No | | | | | | | Ref. | | |
| Yes | | | | | | | 3.22 | ( 1.69 - 6.12 ) | **<0.001** |
| **2) HIV-related characteristics** | | | | | | | | | |
| **HIV status** | | | | | | | | | |
| Negative | | | | | | | Ref. | | |
| Positive | | | | | | | 0.97 | ( 0.40 - 2.31 ) | 0.938 |
| **Partner's HIV status** | | | | | | | | | |
| Negative | | | | | | | Ref. | | |
| Positive | | | | | | | 2.04 | ( 0.82 - 5.09 ) | 0.128 |
| Don't know | | | | | | | 1.04 | ( 0.60 - 1.81 ) | 0.887 |
| **Perceived risk for HIV/STIs** | | | | | | | | | |
| No risk at all | | | | | | | Ref. | | |
| Small | | | | | | | 0.68 | ( 0.39 - 1.20 ) | 0.187 |
| Moderate | | | | | | | 0.79 | ( 0.44 - 1.42 ) | 0.437 |
| Great | | | | | | | 0.77 | ( 0.41 - 1.47 ) | 0.429 |
| **3) HEC use** | | | | | | | | | |
| **Type of HECs** | | | | | | | | | |
| Injectables | | | | | | | Ref. | | |
| Implants | | | | | | | 0.86 | ( 0.56 - 1.31 ) | 0.483 |
| IUD | | | | | | | 1.23 | ( 0.67 - 2.25 ) | 0.511 |
| Pill | | | | | | | 1.36 | ( 0.66 - 2.80 ) | 0.408 |
| Female sterilization | | | | | | | | | perfect success |
| **4) Other psychosocial characterlocalicts** | | | | | | | | | |
| **HIV-related knowledge (HIV-KQ-18)** | | | | | | | 1.04 | ( 0.96 - 1.12 ) | 0.312 |
| **Condom use self-efficacy scale** | | | | | | | 1.03 | ( 1.00 - 1.05 ) | **0.029** |
| **Sexual Relationship Power Scale** | | | | | | | | | |
| Low | | | | | | | Ref. | | |
| Medium | | | | | | | 1.29 | ( 0.81 - 2.05 ) | 0.290 |
| High | | | | | | | 1.42 | ( 0.87 - 2.31 ) | 0.165 |

OR: odds ratio; AOR: adjusted odds ratio; SD: standard deviation; HIV: human immunodeficiency virus; STI: sexually transmitted infection; HEC: highly effective contraceptive; IUD: intrauterine device

a. Adjusted for the cluster effect

b. Adjusted for age, education, religion, wealth index, number of children, pregnancy intention, partner's pregnancy intention, history of unintended pregnancy, multiple sex partnership, HIV status, partner's HIV status, risk perception of HIV/STIs, HIV-related knowledge, condom use self-efficacy, and sexual relationship control power.

c. Intervention*time represents the status of the intervention group at follow-up in comparison with the control group at the baseline.

**Supplementary Table S7. Effects of intervention on consistent dual-method use amonng women at 2 months after enrollement**

| Variables | Model 1 | | | Model 2 | | | Model 3 | | |
|---|---|---|---|---|---|---|---|---|---|
| | OR | (95% CI) | p-value | AOR[a] | (95% CI) | p-value | AOR[b] | (95% CI) | p-value |
| **Intervention** | | | | | | | | | |
| Control | Ref. | | | Ref. | | | Ref. | | |
| Intervention | 11.98 | ( 4.74 - 30.29 ) | **<0.001** | 13.71 | ( 3.59 - 52.43 ) | **<0.001** | 14.53 | ( 3.63 - 58.134 ) | **<0.001** |
| **1) Socio-demographic characteristics** | | | | | | | | | |
| **Age in years** | | | | | | | 1.01 | ( 0.94 - 1.076 ) | 0.856 |
| **Education** | | | | | | | | | |
| Never | | | | | | | Ref. | | |
| Primary and more | | | | | | | 0.69 | ( 0.34 - 1.3896 ) | 0.298 |
| **Religion** | | | | | | | | | |
| Christian | | | | | | | Ref. | | |
| Muslim | | | | | | | 0.93 | ( 0.30 - 2.8508 ) | 0.898 |
| **Wealth index** | | | | | | | | | |
| Poor | | | | | | | Ref. | | |
| Middle | | | | | | | 1.47 | ( 0.70 - 3.1084 ) | 0.307 |
| Rich | | | | | | | 1.37 | ( 0.61 - 3.0864 ) | 0.441 |
| **No. of children** | | | | | | | 0.89 | ( 0.69 - 1.1615 ) | 0.396 |
| **Pregnancy intention** | | | | | | | | | |
| No | | | | | | | Ref. | | |
| Yes | | | | | | | 0.56 | ( 0.20 - 1.5436 ) | 0.264 |
| Don't know | | | | | | | 1.51 | ( 0.39 - 5.8358 ) | 0.551 |
| **Partner's pregnancy intention** | | | | | | | | | |
| No | | | | | | | Ref. | | |
| Yes | | | | | | | 0.89 | ( 0.30 - 2.6386 ) | 0.834 |
| Don't know | | | | | | | 0.59 | ( 0.17 - 2.0459 ) | 0.405 |
| **History of unintended pregnancy** | | | | | | | | | |
| No | | | | | | | Ref. | | |
| Yes | | | | | | | 0.76 | ( 0.39 - 1.4826 ) | 0.421 |
| **Multiple sex partners** | | | | | | | | | |
| No | | | | | | | Ref. | | |
| Yes | | | | | | | 3.21 | ( 1.06 - 9.6705 ) | **0.039** |
| **2) HIV-related characteristics** | | | | | | | | | |
| **HIV status** | | | | | | | | | |
| Negative | | | | | | | Ref. | | |
| Positive | | | | | | | 1.47 | ( 0.39 - 5.5239 ) | 0.566 |
| **Partner's HIV status** | | | | | | | | | |
| Negative | | | | | | | Ref. | | |
| Positive | | | | | | | 1.23 | ( 0.28 - 5.4303 ) | 0.785 |
| Don't know | | | | | | | 1.15 | ( 0.48 - 2.7661 ) | 0.747 |
| **Perceived risk for HIV/STIs** | | | | | | | | | |
| No risk at all | | | | | | | Ref. | | |
| Small | | | | | | | 1.98 | ( 0.57 - 6.9055 ) | 0.283 |
| Moderate | | | | | | | 2.37 | ( 0.67 - 8.43 ) | 0.181 |
| Great | | | | | | | 4.04 | ( 1.10 - 14.824 ) | 0.035 |
| **3) HEC use** | | | | | | | | | |
| **Type of HECs** | | | | | | | | | |
| Injectables | | | | | | | Ref. | | |
| Implants | | | | | | | 0.53 | ( 0.27 - 1.0388 ) | 0.064 |
| IUD | | | | | | | 0.47 | ( 0.14 - 1.5696 ) | 0.219 |
| Pill | | | | | | | 0.16 | ( 0.02 - 1.3684 ) | 0.093 |
| Female sterilization | | | | | | | | | perfect success |
| **4) Other psychosocial charactericts** | | | | | | | | | |
| **HIV-related knowledge (HIV-KQ-18)** | | | | | | | 1.02 | ( 0.90 - 1.156 ) | 0.722 |
| **Condom use self-efficacy scale** | | | | | | | 0.98 | ( 0.94 - 1.0221 ) | 0.359 |
| **Sexual Relationship Power Scale** | | | | | | | | | |
| Low | | | | | | | Ref. | | |
| Medium | | | | | | | 1.36 | ( 0.62 - 2.9545 ) | 0.445 |
| High | | | | | | | 1.87 | ( 0.84 - 4.1729 ) | 0.124 |

OR: odds ratio; AOR: adjusted odds ratio; SD: standard deviation; HIV: human immunodeficiency virus; STI: sexually transmitted infection; HEC: highly effective contraceptive; IUD: intrauterine device

a. Adjusted for the cluster effect

b. Adjusted for age, education, religion, wealth index, number of children, pregnancy intention, partner's pregnancy intention, history of unintended pregnancy, multiple sex partnership, HIV status, partner's HIV status, risk perception of HIV/STIs, HIV-related knowledge, condom use self-efficacy, and sexual relationship control power.

c. Intervention*time represents the status of the intervention group at follow-up in comparison with the control group at the baseline.

**Supplementary Table S8. Effects of intervention on consistent dual-method use amonng women at 4 months after enrollement**

| Variables | Model 1 | | | Model 2 | | | Model 3 | | |
|---|---|---|---|---|---|---|---|---|---|
| | OR | (95% CI) | p-value | AOR[a] | (95% CI) | p-value | AOR[b] | (95% CI) | p-value |
| **Intervention** | | | | | | | | | |
| Control | Ref. | | | Ref. | | | Ref. | | |
| Intervention | 5.22 | ( 2.42 - 11.279 ) | **<0.001** | 6.28 | ( 2.01 - 19.60 ) | **0.002** | 6.30 | ( 2.20 - 18.03 ) | **0.001** |
| **1) Socio-demographic characteristics** | | | | | | | | | |
| **Age in years** | | | | | | | 1.05 | ( 0.98 - 1.12 ) | 0.181 |
| **Education** | | | | | | | | | |
| Never | | | | | | | Ref. | | |
| Primary and more | | | | | | | 0.56 | ( 0.28 - 1.13 ) | 0.104 |
| **Religion** | | | | | | | | | |
| Christian | | | | | | | Ref. | | |
| Muslim | | | | | | | 0.73 | ( 0.19 - 2.82 ) | 0.651 |
| **Wealth index** | | | | | | | | | |
| Poor | | | | | | | Ref. | | |
| Middle | | | | | | | 1.63 | ( 0.74 - 3.56 ) | 0.224 |
| Rich | | | | | | | 1.51 | ( 0.65 - 3.54 ) | 0.340 |
| **No. of children** | | | | | | | 0.79 | ( 0.59 - 1.05 ) | 0.104 |
| **Pregnancy intention** | | | | | | | | | |
| No | | | | | | | Ref. | | |
| Yes | | | | | | | 0.37 | ( 0.13 - 1.06 ) | 0.063 |
| Don't know | | | | | | | 0.61 | ( 0.16 - 2.43 ) | 0.488 |
| **Partner's pregnancy intention** | | | | | | | | | |
| No | | | | | | | Ref. | | |
| Yes | | | | | | | 1.45 | ( 0.47 - 4.49 ) | 0.523 |
| Don't know | | | | | | | 1.08 | ( 0.32 - 3.67 ) | 0.907 |
| **History of unintended pregnancy** | | | | | | | | | |
| No | | | | | | | Ref. | | |
| Yes | | | | | | | 0.52 | ( 0.24 - 1.12 ) | 0.094 |
| **Multiple sex partners** | | | | | | | | | |
| No | | | | | | | Ref. | | |
| Yes | | | | | | | 0.37 | ( 0.05 - 3.05 ) | 0.356 |
| **2) HIV-related characteristics** | | | | | | | | | |
| **HIV status** | | | | | | | | | |
| Negative | | | | | | | Ref. | | |
| Positive | | | | | | | 1.01 | ( 0.24 - 4.31 ) | 0.985 |
| **Partner's HIV status** | | | | | | | | | |
| Negative | | | | | | | Ref. | | |
| Positive | | | | | | | 1.84 | ( 0.36 - 9.30 ) | 0.462 |
| Don't know | | | | | | | 1.63 | ( 0.68 - 3.92 ) | 0.275 |
| **Perceived risk for HIV/STIs** | | | | | | | | | |
| No risk at all | | | | | | | Ref. | | |
| Small | | | | | | | 2.14 | ( 0.58 - 7.93 ) | 0.253 |
| Moderate | | | | | | | 2.15 | ( 0.56 - 8.32 ) | 0.268 |
| Great | | | | | | | 1.65 | ( 0.39 - 7.03 ) | 0.499 |
| **3) HEC use** | | | | | | | | | |
| **Type of HECs** | | | | | | | | | |
| Injectables | | | | | | | Ref. | | |
| Implants | | | | | | | 1.01 | ( 0.49 - 2.06 ) | 0.987 |
| IUD | | | | | | | 1.58 | ( 0.54 - 4.62 ) | 0.400 |
| Pill | | | | | | | 0.66 | ( 0.13 - 3.32 ) | 0.618 |
| Female sterilization | | | | | | | | | perfect success |
| **4) Other psychosocial charactericts** | | | | | | | | | |
| **HIV-related knowledge (HIV-KQ-18)** | | | | | | | 0.72 | ( 0.32 - 1.63 ) | 0.434 |
| **Condom use self-efficacy scale** | | | | | | | 0.96 | ( 0.42 - 2.21 ) | 0.932 |
| **Sexual Relationship Power Scale** | | | | | | | | | |
| Low | | | | | | | Ref. | | |
| Medium | | | | | | | 1.36 | ( 0.62 - 2.9545 ) | 0.445 |
| High | | | | | | | 1.87 | ( 0.84 - 4.1729 ) | 0.124 |

OR: odds ratio; AOR: adjusted odds ratio; SD: standard deviation; HIV: human immunodeficiency virus; STI: sexually transmitted infection; HEC: highly effective contraceptive; IUD: intrauterine device

a. Adjusted for the cluster effect

b. Adjusted for age, education, religion, wealth index, number of children, pregnancy intention, partner's pregnancy intention, history of unintended pregnancy, multiple sex partnership, HIV status, partner's HIV status, risk perception of HIV/STIs, HIV-related knowledge, condom use self-efficacy, and sexual relationship control power.

c. Intervention*time represents the status of the intervention group at follow-up in comparison with the control group at the baseline.

**Supplementary Table S9. Effects of intervention on consistent dual-method use amonng women at 6 months after enrollement**

| Variables | Model 1 | | | Model 2 | | | Model 3 | | |
|---|---|---|---|---|---|---|---|---|---|
| | OR | (95% CI) | p-value | AOR[a] | (95% CI) | p-value | AOR[b] | (95% CI) | p-value |
| **Intervention** | | | | | | | | | |
| Control | Ref. | | | Ref. | | | Ref. | | |
| Intervention | 6.58 | ( 2.53 - 17.074 ) | **<0.001** | 7.80 | ( 1.22 - 49.73 ) | **0.030** | 8.04 | ( 1.17 - 55.08 ) | **0.034** |
| **1) Socio-demographic characteristics** | | | | | | | | | |
| **Age in years** | | | | | | | 1.05 | ( 0.96 - 1.15 ) | 0.311 |
| **Education** | | | | | | | | | |
| Never | | | | | | | Ref. | | |
| Primary and more | | | | | | | 1.18 | ( 0.45 - 3.13 ) | 0.738 |
| **Religion** | | | | | | | | | |
| Christian | | | | | | | Ref. | | |
| Muslim | | | | | | | 1.75 | ( 0.46 - 6.61 ) | 0.409 |
| **Wealth index** | | | | | | | | | |
| Poor | | | | | | | Ref. | | |
| Middle | | | | | | | 1.35 | ( 0.53 - 3.47 ) | 0.528 |
| Rich | | | | | | | 0.75 | ( 0.26 - 2.20 ) | 0.604 |
| **No. of children** | | | | | | | 0.90 | ( 0.64 - 1.28 ) | 0.560 |
| **Pregnancy intention** | | | | | | | | | |
| No | | | | | | | Ref. | | |
| Yes | | | | | | | 0.73 | ( 0.18 - 2.92 ) | 0.657 |
| Don't know | | | | | | | 1.31 | ( 0.23 - 7.51 ) | 0.763 |
| **Partner's pregnancy intention** | | | | | | | | | |
| No | | | | | | | Ref. | | |
| Yes | | | | | | | 1.27 | ( 0.30 - 5.40 ) | 0.743 |
| Don't know | | | | | | | 0.84 | ( 0.17 - 4.17 ) | 0.836 |
| **History of unintended pregnancy** | | | | | | | | | |
| No | | | | | | | Ref. | | |
| Yes | | | | | | | 0.79 | ( 0.32 - 1.96 ) | 0.607 |
| **Multiple sex partners** | | | | | | | | | |
| No | | | | | | | Ref. | | |
| Yes | | | | | | | 1.59 | ( 0.29 - 8.78 ) | 0.597 |
| **2) HIV-related characteristics** | | | | | | | | | |
| **HIV status** | | | | | | | | | |
| Negative | | | | | | | Ref. | | |
| Positive | | | | | | | 4.08 | ( 0.86 - 19.27 ) | 0.076 |
| **Partner's HIV status** | | | | | | | | | |
| Negative | | | | | | | Ref. | | |
| Positive | | | | | | | 0.51 | ( 0.07 - 3.47 ) | 0.489 |
| Don't know | | | | | | | 0.93 | ( 0.30 - 2.92 ) | 0.901 |
| **Perceived risk for HIV/STIs** | | | | | | | | | |
| No risk at all | | | | | | | Ref. | | |
| Small | | | | | | | 1.21 | ( 0.29 - 5.09 ) | 0.791 |
| Moderate | | | | | | | 0.91 | ( 0.20 - 4.25 ) | 0.907 |
| Great | | | | | | | 0.98 | ( 0.20 - 4.82 ) | 0.983 |
| **3) HEC use** | | | | | | | | | |
| **Type of HECs** | | | | | | | | | |
| Injectables | | | | | | | Ref. | | |
| Implants | | | | | | | 1.09 | ( 0.44 - 2.67 ) | 0.853 |
| IUD | | | | | | | 1.95 | ( 0.55 - 6.93 ) | 0.304 |
| Pill | | | | | | | 1.51 | ( 0.27 - 8.57 ) | 0.642 |
| Female sterilization | | | | | | | | | perfect success |
| **4) Other psychosocial charactericts** | | | | | | | | | |
| **HIV-related knowledge (HIV-KQ-18)** | | | | | | | 1.02 | ( 0.87 - 1.18 ) | 0.834 |
| **Condom use self-efficacy scale** | | | | | | | 1.02 | ( 0.97 - 1.07 ) | 0.549 |
| **Sexual Relationship Power Scale** | | | | | | | | | |
| Low | | | | | | | Ref. | | |
| Medium | | | | | | | 0.93 | ( 0.36 - 2.43 ) | 0.885 |
| High | | | | | | | 0.56 | ( 0.18 - 1.71 ) | 0.310 |

OR: odds ratio; AOR: adjusted odds ratio; SD: standard deviation; HIV: human immunodeficiency virus; STI: sexually transmitted infection; HEC: highly effective contraceptive; IUD: intrauterine device

a. Adjusted for the cluster effect

b. Adjusted for age, education, religion, wealth index, number of children, pregnancy intention, partner's pregnancy intention, history of unintended pregnancy, multiple sex partnership, HIV status, partner's HIV status, risk perception of HIV/STIs, HIV-related knowledge, condom use self-efficacy, and sexual relationship control power.

c. Intervention*time represents the status of the intervention group at follow-up in comparison with the control group at the baseline.

**Supplementary Table S10. Effects of intervention on consistent dual-method use amonng women at 8 months after enrollement**

| Variables | Model 1 | | | Model 2 | | | Model 3 | | |
|---|---|---|---|---|---|---|---|---|---|
| | OR | (95% CI) | p-value | AOR[a] | (95% CI) | p-value | AOR[b] | (95% CI) | p-value |
| **Intervention** | | | | | | | | | |
| Control | Ref. | | | Ref. | | | Ref. | | |
| Intervention | 9.43 | ( 3.70 - 24.055 ) | **<0.001** | 9.97 | ( 2.11 - 47.15 ) | **0.004** | 10.72 | ( 2.03 - 56.64 ) | **0.005** |
| **1) Socio-demographic characteristics** | | | | | | | | | |
| **Age in years** | | | | | | | 1.05 | ( 0.96 - 1.14 ) | 0.270 |
| **Education** | | | | | | | | | |
| Never | | | | | | | Ref. | | |
| Primary and more | | | | | | | 0.90 | ( 0.40 - 2.00 ) | 0.788 |
| **Religion** | | | | | | | | | |
| Christian | | | | | | | Ref. | | |
| Muslim | | | | | | | 1.15 | ( 0.31 - 4.29 ) | 0.832 |
| **Wealth index** | | | | | | | | | |
| Poor | | | | | | | Ref. | | |
| Middle | | | | | | | 1.46 | ( 0.63 - 3.38 ) | 0.373 |
| Rich | | | | | | | 1.52 | ( 0.61 - 3.80 ) | 0.373 |
| **No. of children** | | | | | | | 0.89 | ( 0.65 - 1.22 ) | 0.463 |
| **Pregnancy intention** | | | | | | | | | |
| No | | | | | | | Ref. | | |
| Yes | | | | | | | 0.40 | ( 0.12 - 1.34 ) | 0.137 |
| Don't know | | | | | | | 0.93 | ( 0.22 - 3.98 ) | 0.923 |
| **Partner's pregnancy intention** | | | | | | | | | |
| No | | | | | | | Ref. | | |
| Yes | | | | | | | 1.80 | ( 0.47 - 6.86 ) | 0.390 |
| Don't know | | | | | | | 1.34 | ( 0.34 - 5.24 ) | 0.674 |
| **History of unintended pregnancy** | | | | | | | | | |
| No | | | | | | | Ref. | | |
| Yes | | | | | | | 0.86 | ( 0.40 - 1.83 ) | 0.688 |
| **Multiple sex partners** | | | | | | | | | |
| No | | | | | | | Ref. | | |
| Yes | | | | | | | 0.94 | ( 0.17 - 5.16 ) | 0.942 |
| **2) HIV-related characteristics** | | | | | | | | | |
| **HIV status** | | | | | | | | | |
| Negative | | | | | | | Ref. | | |
| Positive | | | | | | | 1.16 | ( 0.20 - 6.63 ) | 0.868 |
| **Partner's HIV status** | | | | | | | | | |
| Negative | | | | | | | Ref. | | |
| Positive | | | | | | | 1.12 | ( 0.18 - 7.00 ) | 0.905 |
| Don't know | | | | | | | 0.41 | ( 0.12 - 1.36 ) | 0.146 |
| **Perceived risk for HIV/STIs** | | | | | | | | | |
| No risk at all | | | | | | | Ref. | | |
| Small | | | | | | | 0.85 | ( 0.27 - 2.70 ) | 0.782 |
| Moderate | | | | | | | 0.96 | ( 0.29 - 3.16 ) | 0.944 |
| Great | | | | | | | 1.20 | ( 0.33 - 4.34 ) | 0.785 |
| **3) HEC use** | | | | | | | | | |
| **Type of HECs** | | | | | | | | | |
| Injectables | | | | | | | Ref. | | |
| Implants | | | | | | | 0.85 | ( 0.38 - 1.89 ) | 0.685 |
| IUD | | | | | | | 2.55 | ( 0.87 - 7.46 ) | 0.087 |
| Pill | | | | | | | 0.60 | ( 0.11 - 3.37 ) | 0.566 |
| Female sterilization | | | | | | | | | perfect success |
| **4) Other psychosocial charactericts** | | | | | | | | | |
| **HIV-related knowledge (HIV-KQ-18)** | | | | | | | 1.02 | ( 0.89 - 1.16 ) | 0.779 |
| **Condom use self-efficacy scale** | | | | | | | 1.00 | ( 0.96 - 1.05 ) | 0.858 |
| **Sexual Relationship Power Scale** | | | | | | | | | |
| Low | | | | | | | Ref. | | |
| Medium | | | | | | | 0.90 | ( 0.37 - 2.16 ) | 0.806 |
| High | | | | | | | 1.11 | ( 0.44 - 2.83 ) | 0.829 |

OR: odds ratio; AOR: adjusted odds ratio; SD: standard deviation; HIV: human immunodeficiency virus; STI: sexually transmitted infection; HEC: highly effective contraceptive; IUD: intrauterine device

a. Adjusted for the cluster effect

b. Adjusted for age, education, religion, wealth index, number of children, pregnancy intention, partner's pregnancy intention, history of unintended pregnancy, multiple sex partnership, HIV status, partner's HIV status, risk perception of HIV/STIs, HIV-related knowledge, condom use self-efficacy, and sexual relationship control power.

c. Intervention*time represents the status of the intervention group at follow-up in comparison with the control group at the baseline.

**Supplementary Table S11. Effects of intervention on communication about HIV/STI risk amonng women at 2 months after enrollement**

| Variables | Model 1 | | | Model 2 | | | Model 3 | | |
|---|---|---|---|---|---|---|---|---|---|
| | OR | (95% CI) | p-value | AOR[a] | (95% CI) | p-value | AOR[b] | (95% CI) | p-value |
| **Intervention** | | | | | | | | | |
| Control | Ref. | | | Ref. | | | Ref. | | |
| Intervention | 2.73 | ( 1.92 - 3.90 ) | **<0.001** | 1.03 | ( 0.73 - 1.46 ) | 0.863 | 0.98 | ( 0.68 - 1.42 ) | 0.920 |
| **Time** | | | | 2.19 | ( 1.62 - 2.97 ) | **<0.001** | 2.29 | ( 1.70 - 3.09 ) | **<0.001** |
| **Intervention*time[c]** | | | | 2.70 | ( 1.72 - 4.23 ) | **<0.001** | 2.70 | ( 1.72 - 4.24 ) | **<0.001** |
| **1) Socio-demographic characteristics** | | | | | | | | | |
| **Age in years** | | | | | | | 0.99 | ( 0.97 - 1.02 ) | 0.518 |
| **Education** | | | | | | | | | |
| Never | | | | | | | Ref. | | |
| Primary and more | | | | | | | 0.92 | ( 0.70 - 1.20 ) | 0.527 |
| **Religion** | | | | | | | | | |
| Christian | | | | | | | Ref. | | |
| Muslim | | | | | | | 0.88 | ( 0.59 - 1.32 ) | 0.540 |
| **Wealth index** | | | | | | | | | |
| Poor | | | | | | | Ref. | | |
| Middle | | | | | | | 0.98 | ( 0.75 - 1.29 ) | 0.909 |
| Rich | | | | | | | 0.97 | ( 0.73 - 1.30 ) | 0.852 |
| **No. of children** | | | | | | | 0.92 | ( 0.84 - 1.02 ) | 0.101 |
| **Pregnancy intention** | | | | | | | | | |
| No | | | | | | | Ref. | | |
| Yes | | | | | | | 0.77 | ( 0.52 - 1.13 ) | 0.185 |
| Don't know | | | | | | | 0.75 | ( 0.45 - 1.23 ) | 0.247 |
| **Partner's pregnancy intention** | | | | | | | | | |
| No | | | | | | | Ref. | | |
| Yes | | | | | | | 1.14 | ( 0.74 - 1.75 ) | 0.560 |
| Don't know | | | | | | | 1.13 | ( 0.73 - 1.76 ) | 0.588 |
| **History of unintended pregnancy** | | | | | | | | | |
| No | | | | | | | Ref. | | |
| Yes | | | | | | | 1.30 | ( 1.02 - 1.66 ) | **0.037** |
| **Multiple sex partners** | | | | | | | | | |
| No | | | | | | | Ref. | | |
| Yes | | | | | | | 1.88 | ( 1.12 - 3.17 ) | **0.017** |
| **2) HIV-related characteristics** | | | | | | | | | |
| **HIV status** | | | | | | | | | |
| Negative | | | | | | | Ref. | | |
| Positive | | | | | | | 2.03 | ( 1.06 - 3.89 ) | **0.034** |
| **Partner's HIV status** | | | | | | | | | |
| Negative | | | | | | | Ref. | | |
| Positive | | | | | | | 0.87 | ( 0.44 - 1.72 ) | 0.683 |
| Don't know | | | | | | | 0.90 | ( 0.65 - 1.25 ) | 0.518 |
| **Perceived risk for HIV/STIs** | | | | | | | | | |
| No risk at all | | | | | | | Ref. | | |
| Small | | | | | | | 1.07 | ( 0.77 - 1.48 ) | 0.704 |
| Moderate | | | | | | | 1.10 | ( 0.78 - 1.55 ) | 0.598 |
| Great | | | | | | | 1.04 | ( 0.71 - 1.53 ) | 0.835 |
| **3) HEC use** | | | | | | | | | |
| **Type of HECs** | | | | | | | | | |
| Injectables | | | | | | | Ref. | | |
| Implants | | | | | | | 0.97 | ( 0.76 - 1.24 ) | 0.817 |
| IUD | | | | | | | 1.17 | ( 0.80 - 1.70 ) | 0.421 |
| Pill | | | | | | | 1.07 | ( 0.68 - 1.70 ) | 0.765 |
| Female sterilization | | | | | | | 3.75 | ( 0.61 - 22.98 ) | 0.153 |
| **4) Other psychosocial charactericts** | | | | | | | | | |
| **HIV-related knowledge (HIV-KQ-18)** | | | | | | | 1.04 | ( 1.00 - 1.09 ) | 0.058 |
| **Condom use self-efficacy scale** | | | | | | | 1.04 | ( 1.02 - 1.05 ) | **<0.001** |
| **Sexual Relationship Power Scale** | | | | | | | | | |
| Low | | | | | | | Ref. | | |
| Medium | | | | | | | 1.02 | ( 0.79 - 1.33 ) | 0.858 |
| High | | | | | | | 1.19 | ( 0.89 - 1.59 ) | 0.248 |

OR: odds ratio; AOR: adjusted odds ratio; SD: standard deviation; HIV: human immunodeficiency virus; STI: sexually transmitted infection; HEC: highly effective contraceptive; IUD: intrauterine device

a. Adjusted for the cluster effect

b. Adjusted for age, education, religion, wealth index, number of children, pregnancy intention, partner's pregnancy intention, history of unintended pregnancy, multiple sex partnership, HIV status, partner's HIV status, risk perception of HIV/STIs, HIV-related knowledge, condom use self-efficacy, and sexual relationship control power.

c. Intervention*time represents the status of the intervention group at follow-up in comparison with the control group at the baseline.

**Supplementary Table S12. Effects of intervention on communication about HIV/STI risk amonng women at 4 months after enrollement**

| Variables | Model 1 | | | Model 2 | | | Model 3 | | |
|---|---|---|---|---|---|---|---|---|---|
| | OR | (95% CI) | p-value | AOR[a] | (95% CI) | p-value | AOR[b] | (95% CI) | p-value |
| **Intervention** | | | | | | | | | |
| Control | Ref. | | | Ref. | | | Ref. | | |
| Intervention | 1.81 | (1.21 - 2.71) | **0.004** | 1.04 | (0.72 - 1.50) | 0.841 | 0.99 | (0.68 - 1.44) | 0.943 |
| **Time** | | | | 5.07 | (3.55 - 7.25) | **<0.001** | 5.72 | (4.08 - 8.02) | **<0.001** |
| **Intervention*time[c]** | | | | 1.76 | (1.08 - 2.86) | **0.023** | 1.76 | (1.07 - 2.89) | **0.025** |
| **1) Socio-demographic characteristics** | | | | | | | | | |
| **Age in years** | | | | | | | 1.00 | (0.98 - 1.03) | 0.973 |
| **Education** | | | | | | | | | |
| Never | | | | | | | Ref. | | |
| Primary and more | | | | | | | 0.88 | (0.66 - 1.17) | 0.372 |
| **Religion** | | | | | | | | | |
| Christian | | | | | | | Ref. | | |
| Muslim | | | | | | | 0.88 | (0.57 - 1.34) | 0.548 |
| **Wealth index** | | | | | | | | | |
| Poor | | | | | | | Ref. | | |
| Middle | | | | | | | 1.01 | (0.76 - 1.34) | 0.947 |
| Rich | | | | | | | 0.88 | (0.64 - 1.19) | 0.396 |
| **No. of children** | | | | | | | 0.92 | (0.83 - 1.02) | 0.109 |
| **Pregnancy intention** | | | | | | | | | |
| No | | | | | | | Ref. | | |
| Yes | | | | | | | 0.97 | (0.64 - 1.45) | 0.868 |
| Don't know | | | | | | | 1.07 | (0.64 - 1.81) | 0.790 |
| **Partner's pregnancy intention** | | | | | | | | | |
| No | | | | | | | Ref. | | |
| Yes | | | | | | | 1.12 | (0.71 - 1.76) | 0.636 |
| Don't know | | | | | | | 0.99 | (0.62 - 1.58) | 0.964 |
| **History of unintended pregnancy** | | | | | | | | | |
| No | | | | | | | Ref. | | |
| Yes | | | | | | | 1.66 | (1.28 - 2.16) | **<0.001** |
| **Multiple sex partners** | | | | | | | | | |
| No | | | | | | | Ref. | | |
| Yes | | | | | | | 1.86 | (1.08 - 3.19) | **0.025** |
| **2) HIV-related characteristics** | | | | | | | | | |
| **HIV status** | | | | | | | | | |
| Negative | | | | | | | Ref. | | |
| Positive | | | | | | | 1.88 | (0.95 - 3.73) | 0.072 |
| **Partner's HIV status** | | | | | | | | | |
| Negative | | | | | | | Ref. | | |
| Positive | | | | | | | 0.96 | (0.46 - 1.98) | 0.907 |
| Don't know | | | | | | | 0.87 | (0.61 - 1.22) | 0.410 |
| **Perceived risk for HIV/STIs** | | | | | | | | | |
| No risk at all | | | | | | | Ref. | | |
| Small | | | | | | | 1.14 | (0.80 - 1.61) | 0.470 |
| Moderate | | | | | | | 1.07 | (0.74 - 1.54) | 0.725 |
| Great | | | | | | | 1.09 | (0.73 - 1.64) | 0.677 |
| **3) HEC use** | | | | | | | | | |
| **Type of HECs** | | | | | | | | | |
| Injectables | | | | | | | Ref. | | |
| Implants | | | | | | | 0.87 | (0.67 - 1.13) | 0.287 |
| IUD | | | | | | | 1.29 | (0.87 - 1.93) | 0.206 |
| Pill | | | | | | | 1.24 | (0.75 - 2.04) | 0.400 |
| Female sterilization | | | | | | | 3.41 | (0.52 - 22.20) | 0.200 |
| **4) Other psychosocial charactericts** | | | | | | | | | |
| **HIV-related knowledge (HIV-KQ-18)** | | | | | | | 1.03 | (0.98 - 1.08) | 0.219 |
| **Condom use self-efficacy scale** | | | | | | | 1.04 | (1.03 - 1.06) | **<0.001** |
| **Sexual Relationship Power Scale** | | | | | | | | | |
| Low | | | | | | | Ref. | | |
| Medium | | | | | | | 1.12 | (0.85 - 1.47) | 0.419 |
| High | | | | | | | 1.32 | (0.97 - 1.81) | 0.075 |

OR: odds ratio; AOR: adjusted odds ratio; SD: standard deviation; HIV: human immunodeficiency virus; STI: sexually transmitted infection; HEC: highly effective contraceptive; IUD: intrauterine device

a. Adjusted for the cluster effect

b. Adjusted for age, education, religion, wealth index, number of children, pregnancy intention, partner's pregnancy intention, history of unintended pregnancy, multiple sex partnership, HIV status, partner's HIV status, risk perception of HIV/STIs, HIV-related knowledge, condom use self-efficacy, and sexual relationship control power.

c. Intervention*time represents the status of the intervention group at follow-up in comparison with the control group at the baseline.

**Supplementary Table S13. Effects of intervention on communication about HIV/STI risk amonng women at 6 months after enrollement**

| Variables | Model 1 | | | Model 2 | | | Model 3 | | |
|---|---|---|---|---|---|---|---|---|---|
| | OR | (95% CI) | p-value | AOR[a] | (95% CI) | p-value | AOR[b] | (95% CI) | p-value |
| **Intervention** | | | | | | | | | |
| Control | Ref. | | | Ref. | | | Ref. | | |
| Intervention | 3.33 | ( 2.13 - 5.20 ) | **<0.001** | 1.05 | ( 0.78 - 1.41 ) | 0.741 | 1.00 | ( 0.74 - 1.35 ) | 0.991 |
| **Time** | | | | 4.12 | ( 3.04 - 5.59 ) | **<0.001** | 4.45 | ( 3.25 - 6.10 ) | **<0.001** |
| **Intervention*time[c]** | | | | 3.23 | ( 1.93 - 5.41 ) | **<0.001** | 3.35 | ( 1.99 - 5.66 ) | **<0.001** |
| **1) Socio-demographic characteristics** | | | | | | | | | |
| **Age in years** | | | | | | | 0.99 | ( 0.96 - 1.01 ) | 0.298 |
| **Education** | | | | | | | | | |
| Never | | | | | | | Ref. | | |
| Primary and more | | | | | | | 0.91 | ( 0.69 - 1.20 ) | 0.492 |
| **Religion** | | | | | | | | | |
| Christian | | | | | | | Ref. | | |
| Muslim | | | | | | | 0.99 | ( 0.65 - 1.51 ) | 0.967 |
| **Wealth index** | | | | | | | | | |
| Poor | | | | | | | Ref. | | |
| Middle | | | | | | | 1.01 | ( 0.76 - 1.34 ) | 0.958 |
| Rich | | | | | | | 0.90 | ( 0.66 - 1.22 ) | 0.481 |
| **No. of children** | | | | | | | 0.95 | ( 0.86 - 1.05 ) | 0.291 |
| **Pregnancy intention** | | | | | | | | | |
| No | | | | | | | Ref. | | |
| Yes | | | | | | | 1.12 | ( 0.75 - 1.67 ) | 0.576 |
| Don't know | | | | | | | 0.93 | ( 0.56 - 1.56 ) | 0.795 |
| **Partner's pregnancy intention** | | | | | | | | | |
| No | | | | | | | Ref. | | |
| Yes | | | | | | | 0.91 | ( 0.58 - 1.42 ) | 0.678 |
| Don't know | | | | | | | 0.89 | ( 0.56 - 1.41 ) | 0.621 |
| **History of unintended pregnancy** | | | | | | | | | |
| No | | | | | | | Ref. | | |
| Yes | | | | | | | 1.45 | ( 1.12 - 1.87 ) | **0.005** |
| **Multiple sex partners** | | | | | | | | | |
| No | | | | | | | Ref. | | |
| Yes | | | | | | | 2.29 | ( 1.32 - 3.99 ) | **0.003** |
| **2) HIV-related characteristics** | | | | | | | | | |
| **HIV status** | | | | | | | | | |
| Negative | | | | | | | Ref. | | |
| Positive | | | | | | | 1.19 | ( 0.62 - 2.29 ) | 0.591 |
| **Partner's HIV status** | | | | | | | | | |
| Negative | | | | | | | Ref. | | |
| Positive | | | | | | | 1.09 | ( 0.54 - 2.19 ) | 0.807 |
| Don't know | | | | | | | 1.03 | ( 0.73 - 1.45 ) | 0.858 |
| **Perceived risk for HIV/STIs** | | | | | | | | | |
| No risk at all | | | | | | | Ref. | | |
| Small | | | | | | | 1.14 | ( 0.81 - 1.61 ) | 0.443 |
| Moderate | | | | | | | 1.03 | ( 0.72 - 1.48 ) | 0.875 |
| Great | | | | | | | 1.05 | ( 0.70 - 1.56 ) | 0.828 |
| **3) HEC use** | | | | | | | | | |
| **Type of HECs** | | | | | | | | | |
| Injectables | | | | | | | Ref. | | |
| Implants | | | | | | | 1.03 | ( 0.80 - 1.33 ) | 0.810 |
| IUD | | | | | | | 0.97 | ( 0.66 - 1.42 ) | 0.869 |
| Pill | | | | | | | 1.30 | ( 0.80 - 2.12 ) | 0.288 |
| Female sterilization | | | | | | | 1.51 | ( 0.25 - 9.05 ) | 0.650 |
| **4) Other psychosocial charactericts** | | | | | | | | | |
| **HIV-related knowledge (HIV-KQ-18)** | | | | | | | 1.04 | ( 0.99 - 1.08 ) | 0.106 |
| **Condom use self-efficacy scale** | | | | | | | 1.03 | ( 1.02 - 1.05 ) | **<0.001** |
| **Sexual Relationship Power Scale** | | | | | | | | | |
| Low | | | | | | | Ref. | | |
| Medium | | | | | | | 1.09 | ( 0.83 - 1.43 ) | 0.551 |
| High | | | | | | | 1.27 | ( 0.94 - 1.72 ) | 0.122 |

OR: odds ratio; AOR: adjusted odds ratio; SD: standard deviation; HIV: human immunodeficiency virus; STI: sexually transmitted infection; HEC: highly effective contraceptive; IUD: intrauterine device

a. Adjusted for the cluster effect

b. Adjusted for age, education, religion, wealth index, number of children, pregnancy intention, partner's pregnancy intention, history of unintended pregnancy, multiple sex partnership, HIV status, partner's HIV status, risk perception of HIV/STIs, HIV-related knowledge, condom use self-efficacy, and sexual relationship control power.

c. Intervention*time represents the status of the intervention group at follow-up in comparison with the control group at the baseline.

**Supplementary Table S14. Effects of intervention on communication about HIV/STI risk amonng women at 8 months after enrollement**

| Variables | Model 1 | | | Model 2 | | | Model 3 | | |
|---|---|---|---|---|---|---|---|---|---|
| | OR | (95% CI) | p-value | AOR[a] | (95% CI) | p-value | AOR[b] | (95% CI) | p-value |
| **Intervention** | | | | | | | | | |
| Control | Ref. | | | Ref. | | | Ref. | | |
| Intervention | 1.75 | ( 1.22 - 2.52 ) | **0.002** | 1.04 | ( 0.68 - 1.59 ) | 0.858 | 0.99 | ( 0.65 - 1.51 ) | 0.959 |
| **Time** | | | | 3.65 | ( 2.71 - 4.92 ) | **<0.001** | 3.85 | ( 2.83 - 5.22 ) | **<0.001** |
| **Intervention*time[c]** | | | | 1.75 | ( 1.12 - 2.74 ) | **0.015** | 1.80 | ( 1.14 - 2.84 ) | **0.012** |
| **1) Socio-demographic characteristics** | | | | | | | | | |
| **Age in years** | | | | | | | 0.99 | ( 0.97 - 1.02 ) | 0.555 |
| **Education** | | | | | | | | | |
| Never | | | | | | | Ref. | | |
| Primary and more | | | | | | | 1.07 | ( 0.82 - 1.40 ) | 0.610 |
| **Religion** | | | | | | | | | |
| Christian | | | | | | | Ref. | | |
| Muslim | | | | | | | 0.85 | ( 0.57 - 1.29 ) | 0.453 |
| **Wealth index** | | | | | | | | | |
| Poor | | | | | | | Ref. | | |
| Middle | | | | | | | 1.05 | ( 0.79 - 1.38 ) | 0.749 |
| Rich | | | | | | | 0.98 | ( 0.73 - 1.33 ) | 0.918 |
| **No. of children** | | | | | | | 0.97 | ( 0.88 - 1.07 ) | 0.552 |
| **Pregnancy intention** | | | | | | | | | |
| No | | | | | | | Ref. | | |
| Yes | | | | | | | 0.94 | ( 0.64 - 1.39 ) | 0.763 |
| Don't know | | | | | | | 0.94 | ( 0.57 - 1.55 ) | 0.814 |
| **Partner's pregnancy intention** | | | | | | | | | |
| No | | | | | | | Ref. | | |
| Yes | | | | | | | 1.17 | ( 0.76 - 1.80 ) | 0.474 |
| Don't know | | | | | | | 1.20 | ( 0.77 - 1.87 ) | 0.432 |
| **History of unintended pregnancy** | | | | | | | | | |
| No | | | | | | | Ref. | | |
| Yes | | | | | | | 1.44 | ( 1.13 - 1.85 ) | **0.004** |
| **Multiple sex partners** | | | | | | | | | |
| No | | | | | | | Ref. | | |
| Yes | | | | | | | 1.92 | ( 1.13 - 3.24 ) | **0.015** |
| **2) HIV-related characteristics** | | | | | | | | | |
| **HIV status** | | | | | | | | | |
| Negative | | | | | | | Ref. | | |
| Positive | | | | | | | 2.07 | ( 1.07 - 3.99 ) | **0.031** |
| **Partner's HIV status** | | | | | | | | | |
| Negative | | | | | | | Ref. | | |
| Positive | | | | | | | 0.72 | ( 0.36 - 1.43 ) | 0.345 |
| Don't know | | | | | | | 0.87 | ( 0.62 - 1.21 ) | 0.405 |
| **Perceived risk for HIV/STIs** | | | | | | | | | |
| No risk at all | | | | | | | Ref. | | |
| Small | | | | | | | 1.15 | ( 0.83 - 1.61 ) | 0.398 |
| Moderate | | | | | | | 1.19 | ( 0.84 - 1.69 ) | 0.320 |
| Great | | | | | | | 1.16 | ( 0.79 - 1.72 ) | 0.446 |
| **3) HEC use** | | | | | | | | | |
| **Type of HECs** | | | | | | | | | |
| Injectables | | | | | | | Ref. | | |
| Implants | | | | | | | 1.11 | ( 0.86 - 1.42 ) | 0.426 |
| IUD | | | | | | | 1.18 | ( 0.81 - 1.72 ) | 0.399 |
| Pill | | | | | | | 1.08 | ( 0.68 - 1.72 ) | 0.753 |
| Female sterilization | | | | | | | 2.74 | ( 0.44 - 17.17 ) | 0.283 |
| **4) Other psychosocial charactericts** | | | | | | | | | |
| **HIV-related knowledge (HIV-KQ-18)** | | | | | | | 1.03 | ( 0.99 - 1.08 ) | 0.148 |
| **Condom use self-efficacy scale** | | | | | | | 1.03 | ( 1.01 - 1.04 ) | **<0.001** |
| **Sexual Relationship Power Scale** | | | | | | | | | |
| Low | | | | | | | Ref. | | |
| Medium | | | | | | | 1.28 | ( 0.98 - 1.67 ) | 0.066 |
| High | | | | | | | 1.44 | ( 1.07 - 1.94 ) | **0.016** |

OR: odds ratio; AOR: adjusted odds ratio; SD: standard deviation; HIV: human immunodeficiency virus; STI: sexually transmitted infection; HEC: highly effective contraceptive; IUD: intrauterine device

a. Adjusted for the cluster effect

b. Adjusted for age, education, religion, wealth index, number of children, pregnancy intention, partner's pregnancy intention, history of unintended pregnancy, multiple sex partnership, HIV status, partner's HIV status, risk perception of HIV/STIs, HIV-related knowledge, condom use self-efficacy, and sexual relationship control power.

c. Intervention*time represents the status of the intervention group at follow-up in comparison with the control group at the baseline.

**Supplementary Table S15. Effects of intervention on the incidence of pregnancy amonng women at 2 months after enrollement**

| Variables | Model 1 | | | Model 2 | | | Model 3 | | |
|---|---|---|---|---|---|---|---|---|---|
| | OR | (95% CI) | p-value | AOR[a] | (95% CI) | p-value | AOR[b] | (95% CI) | p-value |
| **Intervention** | | | | | | | | | |
| Control | Ref. | | | Ref. | | | Ref. | | |
| Intervention | 0.69 | ( 0.15 - 3.0905 ) | 0.624 | 0.69 | ( 0.15 - 3.09 ) | **0.002** | 1.27 | ( 0.19 - 8.65 ) | 0.804 |
| **1) Socio-demographic characteristics** | | | | | | | | | |
| **Age in years** | | | | | | | 1.13 | ( 0.93 - 1.38 ) | 0.207 |
| **Education** | | | | | | | | | |
| Never | | | | | | | Ref. | | |
| Primary and more | | | | | | | 1.13 | ( 0.08 - 15.65 ) | 0.925 |
| **Religion** | | | | | | | | | |
| Christian | | | | | | | Ref. | | |
| Muslim | | | | | | | | | perfect success |
| **Wealth index** | | | | | | | | | |
| Poor | | | | | | | Ref. | | |
| Middle | | | | | | | 3.38 | ( 0.17 - 68.99 ) | 0.429 |
| Rich | | | | | | | 3.36 | ( 0.18 - 61.53 ) | 0.414 |
| **No. of children** | | | | | | | 0.59 | ( 0.25 - 1.39 ) | 0.226 |
| **Pregnancy intention** | | | | | | | | | |
| No | | | | | | | Ref. | | |
| Yes | | | | | | | 0.12 | ( 0.00 - 2.76 ) | 0.183 |
| Don't know | | | | | | | | | perfect success |
| **Partner's pregnancy intention** | | | | | | | | | |
| No | | | | | | | Ref. | | |
| Yes | | | | | | | 2.12 | ( 0.09 - 47.51 ) | 0.636 |
| Don't know | | | | | | | | | perfect success |
| **History of unintended pregnancy** | | | | | | | | | |
| No | | | | | | | Ref. | | |
| Yes | | | | | | | 1.15 | ( 0.15 - 8.78 ) | 0.890 |
| **Multiple sex partners** | | | | | | | | | |
| No | | | | | | | Ref. | | |
| Yes | | | | | | | | | collinearity |
| **2) HIV-related characteristics** | | | | | | | | | |
| **HIV status** | | | | | | | | | |
| Negative | | | | | | | Ref. | | |
| Positive | | | | | | | 1.07 | ( 0.00 - 1320.72 ) | 0.986 |
| **Partner's HIV status** | | | | | | | | | |
| Negative | | | | | | | Ref. | | |
| Positive | | | | | | | 6.75 | ( 0.01 - 8258.21 ) | 0.599 |
| Don't know | | | | | | | 3.91 | ( 0.29 - 52.16 ) | 0.302 |
| **Perceived risk for HIV/STIs** | | | | | | | | | |
| No risk at all | | | | | | | Ref. | | |
| Small | | | | | | | 0.93 | ( 0.11 - 8.07 ) | 0.950 |
| Moderate | | | | | | | 0.46 | ( 0.04 - 4.70 ) | 0.512 |
| Great | | | | | | | | | perfect success |
| **3) HEC use** | | | | | | | | | |
| **Type of HECs** | | | | | | | | | |
| Injectables | | | | | | | Ref. | | |
| Implants | | | | | | | 3.55 | ( 0.48 - 26.01 ) | 0.213 |
| IUD | | | | | | | 1.93 | ( 0.11 - 35.13 ) | 0.656 |
| Pill | | | | | | | | | perfect success |
| Female sterilization | | | | | | | | | collinearity |
| **4) Other psychosocial charactericts** | | | | | | | | | |
| **HIV-related knowledge (HIV-KQ-18)** | | | | | | | 0.96 | ( 0.64 - 1.45 ) | 0.843 |
| **Condom use self-efficacy scale** | | | | | | | 0.90 | ( 0.81 - 0.99 ) | **0.039** |
| **Sexual Relationship Power Scale** | | | | | | | | | |
| Low | | | | | | | Ref. | | |
| Medium | | | | | | | 0.52 | ( 0.05 - 5.38 ) | 0.580 |
| High | | | | | | | 3.74 | ( 0.33 - 41.91 ) | 0.285 |

OR: odds ratio; AOR: adjusted odds ratio; SD: standard deviation; HIV: human immunodeficiency virus; STI: sexually transmitted infection; HEC: highly effective contraceptive; IUD: intrauterine device

a. Adjusted for the cluster effect

b. Adjusted for age, education, religion, wealth index, number of children, pregnancy intention, partner's pregnancy intention, history of unintended pregnancy, multiple sex partnership, HIV status, partner's HIV status, risk perception of HIV/STIs, HIV-related knowledge, condom use self-efficacy, and sexual relationship control power.

c. Intervention*time represents the status of the intervention group at follow-up in comparison with the control group at the baseline.

**Supplementary Table S16. Effects of intervention on the incidence of pregnancy amonng women at 4 months after enrollement**

| Variables | Model 1 | | | Model 2 | | | Model 3 | | |
|---|---|---|---|---|---|---|---|---|---|
| | OR | (95% CI) | p-value | AOR[a] | (95% CI) | p-value | AOR[b] | (95% CI) | p-value |
| **Intervention** | | | | | | | | | |
| Control | Ref. | | | Ref. | | | Ref. | | |
| Intervention | 0.47 | ( 0.09 - 2.5689 ) | 0.382 | 0.47 | ( 0.09 - 2.57 ) | 0.382 | 0.24 | ( 0.00 - 16.42 ) | 0.506 |
| **1) Socio-demographic characteristics** | | | | | | | | | |
| **Age in years** | | | | | | | 0.86 | ( 0.54 - 1.35 ) | 0.505 |
| **Education** | | | | | | | | | |
| Never | | | | | | | Ref. | | |
| Primary and more | | | | | | | | | perfect success |
| **Religion** | | | | | | | | | |
| Christian | | | | | | | Ref. | | |
| Muslim | | | | | | | | | perfect success |
| **Wealth index** | | | | | | | | | |
| Poor | | | | | | | Ref. | | |
| Middle | | | | | | | 0.12 | ( 0.00 - 5.29 ) | 0.274 |
| Rich | | | | | | | | | perfect success |
| **No. of children** | | | | | | | 1.06 | ( 0.25 - 4.46 ) | 0.938 |
| **Pregnancy intention** | | | | | | | | | |
| No | | | | | | | Ref. | | |
| Yes | | | | | | | 0.03 | ( 0.00 - 19.06 ) | 0.281 |
| Don't know | | | | | | | 0.05 | ( 0.00 - 23.60 ) | 0.335 |
| **Partner's pregnancy intention** | | | | | | | | | |
| No | | | | | | | Ref. | | |
| Yes | | | | | | | 0.25 | ( 0.00 - 32.44 ) | 0.578 |
| Don't know | | | | | | | | | perfect success |
| **History of unintended pregnancy** | | | | | | | | | |
| No | | | | | | | Ref. | | |
| Yes | | | | | | | 2.37 | ( 0.09 - 65.37 ) | 0.609 |
| **Multiple sex partners** | | | | | | | | | |
| No | | | | | | | Ref. | | |
| Yes | | | | | | | | | perfect success |
| **2) HIV-related characteristics** | | | | | | | | | |
| **HIV status** | | | | | | | | | |
| Negative | | | | | | | Ref. | | |
| Positive | | | | | | | | | perfect success |
| **Partner's HIV status** | | | | | | | | | |
| Negative | | | | | | | Ref. | | |
| Positive | | | | | | | | | perfect success |
| Don't know | | | | | | | | | perfect success |
| **Perceived risk for HIV/STIs** | | | | | | | | | |
| No risk at all | | | | | | | Ref. | | |
| Small | | | | | | | 0.27 | ( 0.01 - 6.67 ) | 0.427 |
| Moderate | | | | | | | 1.01 | ( 0.05 - 20.07 ) | 0.997 |
| Great | | | | | | | | | perfect success |
| **3) HEC use** | | | | | | | | | |
| **Type of HECs** | | | | | | | | | |
| Injectables | | | | | | | Ref. | | |
| Implants | | | | | | | 0.16 | ( 0.01 - 3.19 ) | 0.228 |
| IUD | | | | | | | | | perfect success |
| Pill | | | | | | | 0.93 | ( 0.02 - 43.06 ) | 0.970 |
| Female sterilization | | | | | | | | | collinearity |
| **4) Other psychosocial charactericts** | | | | | | | | | |
| **HIV-related knowledge (HIV-KQ-18)** | | | | | | | 0.87 | ( 0.56 - 1.36 ) | 0.543 |
| **Condom use self-efficacy scale** | | | | | | | 1.06 | ( 0.91 - 1.23 ) | 0.476 |
| **Sexual Relationship Power Scale** | | | | | | | | | |
| Low | | | | | | | Ref. | | |
| Medium | | | | | | | 0.37 | ( 0.01 - 9.78 ) | 0.555 |
| High | | | | | | | 0.75 | ( 0.03 - 17.19 ) | 0.859 |

OR: odds ratio; AOR: adjusted odds ratio; SD: standard deviation; HIV: human immunodeficiency virus; STI: sexually transmitted infection; HEC: highly effective contraceptive; IUD: intrauterine device

a. Adjusted for the cluster effect

b. Adjusted for age, education, religion, wealth index, number of children, pregnancy intention, partner's pregnancy intention, history of unintended pregnancy, multiple sex partnership, HIV status, partner's HIV status, risk perception of HIV/STIs, HIV-related knowledge, condom use self-efficacy, and sexual relationship control power.

c. Intervention*time represents the status of the intervention group at follow-up in comparison with the control group at the baseline.

**Supplementary Table S17. Effects of intervention on the incidence of pregnancy amonng women at 8 months after enrollement**

| Variables | Model 1 | | | Model 2 | | | Model 3 | | |
|---|---|---|---|---|---|---|---|---|---|
| | OR | (95% CI) | p-value | AOR[a] | (95% CI) | p-value | AOR[b] | (95% CI) | p-value |
| **Intervention** | | | | | | | | | |
| Control | Ref. | | | Ref. | | | Ref. | | |
| Intervention | 0.38 | ( 0.07 - 1.9557 ) | 0.246 | 0.38 | ( 0.07 - 2.19 ) | 0.281 | 0.40 | ( 0.02 - 8.19 ) | 0.552 |
| **1) Socio-demographic characteristics** | | | | | | | | | |
| **Age in years** | | | | | | | 0.85 | ( 0.64 - 1.13 ) | 0.263 |
| **Education** | | | | | | | | | |
| Never | | | | | | | Ref. | | |
| Primary and more | | | | | | | | | perfect success |
| **Religion** | | | | | | | | | |
| Christian | | | | | | | Ref. | | |
| Muslim | | | | | | | | | perfect success |
| **Wealth index** | | | | | | | | | |
| Poor | | | | | | | Ref. | | |
| Middle | | | | | | | 0.08 | ( 0.01 - 1.14 ) | 0.062 |
| Rich | | | | | | | | | perfect success |
| **No. of children** | | | | | | | 1.23 | ( 0.40 - 3.83 ) | 0.716 |
| **Pregnancy intention** | | | | | | | | | |
| No | | | | | | | Ref. | | |
| Yes | | | | | | | | | perfect success |
| Don't know | | | | | | | | | perfect success |
| **Partner's pregnancy intention** | | | | | | | | | |
| No | | | | | | | Ref. | | |
| Yes | | | | | | | | | perfect success |
| Don't know | | | | | | | | | perfect success |
| **History of unintended pregnancy** | | | | | | | | | |
| No | | | | | | | Ref. | | |
| Yes | | | | | | | 0.92 | ( 0.05 - 16.15 ) | 0.955 |
| **Multiple sex partners** | | | | | | | | | |
| No | | | | | | | Ref. | | |
| Yes | | | | | | | | | perfect success |
| **2) HIV-related characteristics** | | | | | | | | | |
| **HIV status** | | | | | | | | | |
| Negative | | | | | | | Ref. | | |
| Positive | | | | | | | 6.80 | ( 0.17 - 272.13 ) | 0.309 |
| **Partner's HIV status** | | | | | | | | | |
| Negative | | | | | | | Ref. | | |
| Positive | | | | | | | | | perfect success |
| Don't know | | | | | | | | | perfect success |
| **Perceived risk for HIV/STIs** | | | | | | | | | |
| No risk at all | | | | | | | Ref. | | |
| Small | | | | | | | 2.90 | ( 0.34 - 24.54 ) | 0.328 |
| Moderate | | | | | | | | | collinearity |
| Great | | | | | | | | | perfect success |
| **3) HEC use** | | | | | | | | | |
| **Type of HECs** | | | | | | | | | |
| Injectables | | | | | | | Ref. | | |
| Implants | | | | | | | 0.09 | ( 0.00 - 2.08 ) | 0.133 |
| IUD | | | | | | | | | perfect success |
| Pill | | | | | | | 0.80 | ( 0.04 - 16.43 ) | 0.885 |
| Female sterilization | | | | | | | | | perfect success |
| **4) Other psychosocial charactericts** | | | | | | | | | |
| **HIV-related knowledge (HIV-KQ-18)** | | | | | | | 0.86 | ( 0.56 - 1.34 ) | 0.507 |
| **Condom use self-efficacy scale** | | | | | | | 0.92 | ( 0.79 - 1.07 ) | 0.255 |
| **Sexual Relationship Power Scale** | | | | | | | | | |
| Low | | | | | | | Ref. | | |
| Medium | | | | | | | 2.24 | ( 0.15 - 34.45 ) | 0.564 |
| High | | | | | | | 3.01 | ( 0.08 - 113.51 ) | 0.552 |

OR: odds ratio; AOR: adjusted odds ratio; SD: standard deviation; HIV: human immunodeficiency virus; STI: sexually transmitted infection; HEC: highly effective contraceptive; IUD: intrauterine device

a. Adjusted for the cluster effect

b. Adjusted for age, education, religion, wealth index, number of children, pregnancy intention, partner's pregnancy intention, history of unintended pregnancy, multiple sex partnership, HIV status, partner's HIV status, risk perception of HIV/STIs, HIV-related knowledge, condom use self-efficacy, and sexual relationship control power.

c. Intervention*time represents the status of the intervention group at follow-up in comparison with the control group at the baseline.
